# Neural shape models enable cohort-scale simulation of knee cartilage mechanics

**DOI:** 10.64898/2026.10.29.26366866

**Authors:** Anthony A. Gatti, Kathryn R. Marusich, Allison Clouthier, McKenzie S. White, Carmichael Ong, Amir Esrafilian, Colin Smith, Julie Kolesar, Tian Tan, Scott Uhlrich, Constance Chu, Seth Sherman, Garry E. Gold, Scott L. Delp, Feliks Kogan, Akshay S. Chaudhari

## Abstract

Knee osteoarthritis affects over 350 million people worldwide, is a leading cause of disability, and carries substantial costs. Cartilage mechanical load is believed to play a critical role in osteoarthritis pathophysiology, yet clinical biomechanics research uses external measures of joint loading that do not capture tissue-level load. We developed a fully-automated pipeline that builds biomechanical knee models from a single MRI using deep learning-based segmentation and neural shape models, and elastic foundation contact to simulate gait. Applied to 756 knees from the Osteoarthritis Initiative, each simulated under the same 10 standardized gaits, the pipeline converged in 99.6% of 7,560 simulations. Medial tibial pressure decreased from healthy to incident to mild osteoarthritis as disease-related shape changes enlarged contact area, an effect reproduced in simulations of synthetic knees. Yet within knees, medial cartilage thinned fastest over six years in regions of highest pressure, with stronger relationships in incident and mild osteoarthritis knees. Automated simulations provided cohort-scale, tissue-level support for mechanical theories of osteoarthritis progression and a framework for prospectively quantifying cartilage load to guide mechanical interventions.

## Introduction

Knee osteoarthritis (OA) affects over 350 million people worldwide, is a leading cause of disability and reduced quality of life, and carries billions of dollars in direct and indirect costs^1–4^. Despite this burden, pharmacologic options remain limited to symptom management, without the ability to slow or prevent OA. Disease modifying interventions with the strongest evidence all induce mechanical change: high tibial osteotomy^5^, weight loss^6^, and gait retraining^7^. Yet each intervention is often evaluated with external mechanical surrogates, not the loading cartilage experiences, so treatments cannot be aimed at the tissue at risk.

Due to the hypothesized mechanical basis of OA^8^, gait analysis is a cornerstone in understanding knee OA risk, incidence, and progression. Typical gait studies investigate external measures of joint loading such as the knee adduction or flexion moments^9–12^; the adduction moment is a relative measure of gross medial knee load. Meta-analyses indicate that elevated peak knee adduction moment (KAM) is associated with increased risk of OA progression, though the effect is modest (odds ratio: 1.88)^13^. This is likely because the knee adduction moment is a surrogate for cartilage load, the target tissue of interest. Personalizing mechanical interventions to unload vulnerable cartilage requires directly quantifying cartilage mechanics.

Tissue-level mechanics are quantified by combining human movement data, three-dimensional anatomic models, and computational models of tissue contact, each requiring manual effort that limits studies to small samples. Finite element models (FEM) of contact quantify depth-dependent stress and strain while accounting for cartilage biology such as proteoglycan content and collagen orientation^14,15^. However, a single FEM can take days to weeks to build and simulate. Simplified elastic foundation contact models treat cartilage as a “bed of springs”^16,17^ sacrificing depth-dependent outcomes but enabling simulation of full gait cycles in minutes while still being validated against *in vivo* joint loading^18^ and MRI-measured knee kinematics^19^. These models have been used to test whether muscle can compensate for ligament loss^20^, how altering joint shape changes cartilage pressure^21^, and how virtual surgery changes movement^22^. Even these faster models still rely on anatomy obtained from hours of manual MRI segmentation and hand-tuned model parameters. Thus, to study cartilage mechanics in large cohorts necessary to link loading to OA progression requires automating each step.

Deep learning has largely automated segmentation; in the knee it is well established for bone, cartilage, and menisci^23–27^. However, computational biomechanical models still require smooth anatomic surfaces free of imaging artefacts, ligament and tendon attachment sites, and anatomically corresponding points across knees for data analysis. Neural shape models, deep neural networks that encode each anatomic surface as a compact latent vector similar to principal components, address these gaps^28^. Surfaces fitted with a neural shape model are smooth and high-resolution, and because the model’s latent space is continuous, interpolating between two knees deforms one surface smoothly into the other, carrying every point and every ligament attachment along^29,30^. The same latent space also serves as a coordinate system for anatomy where directions encode disease^28,29^, and generating anatomy along these directions yields synthetic knees that could isolate how disease-related shape alters cartilage load. Encoded in a neural shape model, anatomy is no longer a fixed input to a biomechanical model but a variable that can be personalized, pooled across knees, perturbed systematically, and tested as a predictor in its own right.

Here we develop a fully automated pipeline, with neural shape models at its foundation, that builds personalized biomechanical knee models from a single MRI. Deep learning segmentation extracts anatomic surfaces, neural shape modelling parameterizes them, and the resulting knee model is used in an OpenSim simulation of cartilage contact pressures during gait. We apply this pipeline to baseline MRIs of 756 knees from the Osteoarthritis Initiative (OAI), simulating stance-phase pressures in every knee under the same ten gait cycles, one from a healthy adult and nine from a knee OA clinical trial. Correlating each knee’s simulated cartilage pressures with its own cartilage thinning over the following six years, we perform the first cohort-scale analysis co-localizing tissue-level mechanics with structural OA progression.

Across 756 knees and ten gait cycles, we demonstrate that:

1. Between knees, under identical gait, OA-related bone remodelling enlarges cartilage contact area lowering cartilage pressures.
2. Synthetic knees generated along the healthy-to-mild OA shape axis reproduce this effect, showing systematically increased cartilage surface area and decreasing pressures.
3. Within a knee, medial regions experiencing the highest pressure thin the fastest.
4. Pressure identifies where cartilage thins, but joint shape, not pressure, best discriminates who develops OA (area under the receiver operating characteristic curve [AUROC], shape: 0.69 vs. pressure: 0.56).5. Defining weight-bearing cartilage by simulated contact region rather than anatomical landmarks improves sensitivity to longitudinal femoral cartilage thinning (standardized response mean +0.06 to +0.15).

We release all derived data, custom libraries, and end-to-end pipelines at github.com/gattia/comak_knee_oa_simulation, enabling full reproducibility.

## Results

Our approach builds a personalized biomechanical knee model from a single MRI (**Figure 1**). We extract personalized bones, cartilage, and menisci from MRI using a validated deep learning segmentation model^23^ and fit high resolution three-dimensional anatomic surfaces to these segmentations using deep learning-based neural shape models^28^. We determined personalized ligament attachment sites and wrap surfaces, which route ligaments and tendons over bone, using a novel neural shape model interpolation approach (Methods). The resulting biomechanical knee model, parameterized entirely by latent vectors, is integrated into a validated whole-body OpenSim model^19^ to simulate cartilage contact during gait^18,20^.

**Figure 1.**
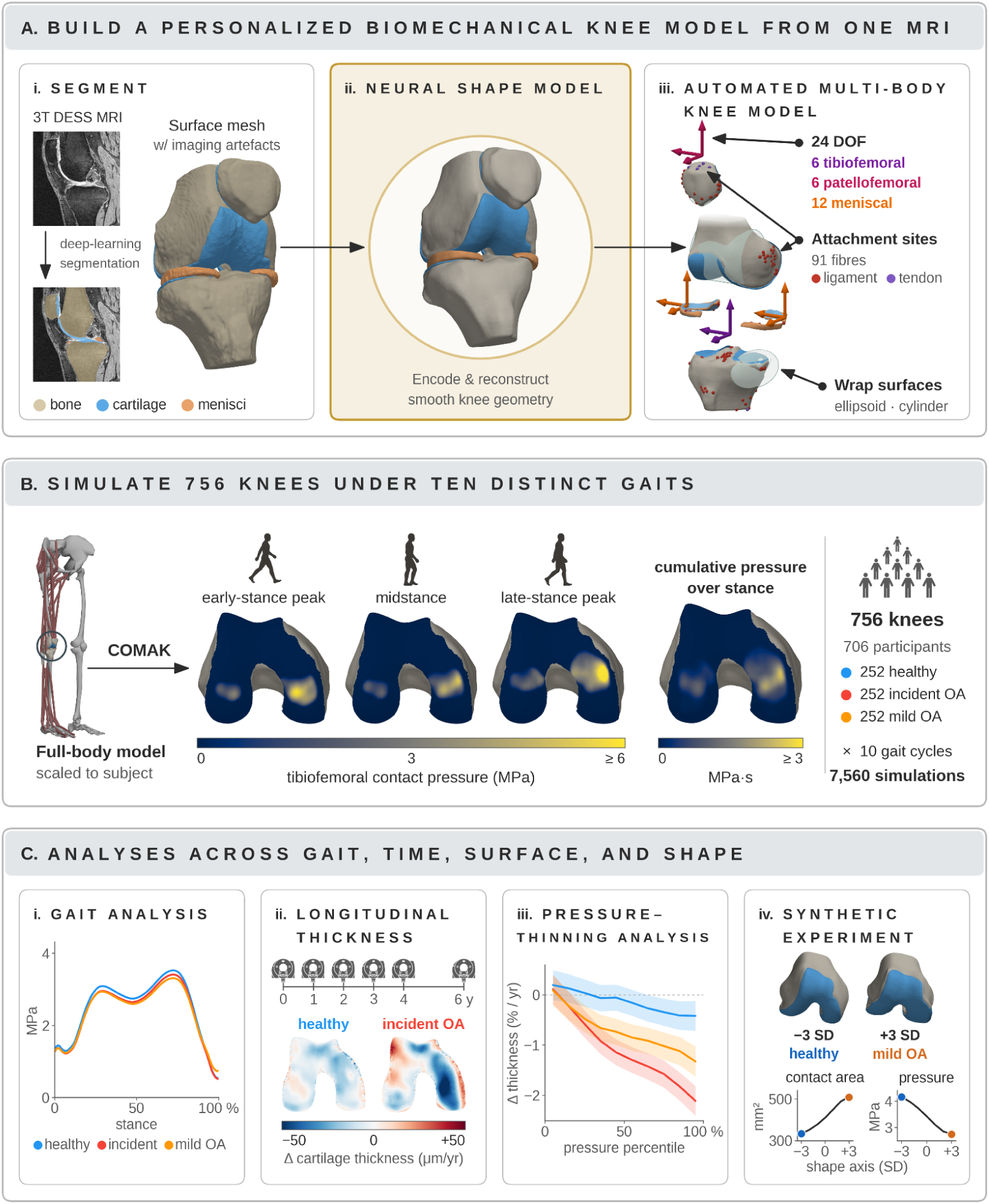
Visualization of the overarching automated pipeline (A, B) and data analyses (C). **A.i.** A 3D double echo in steady state (DESS) knee MRI is segmented using a convolutional neural network, and the resulting segmentation is post-processed to extract bone, cartilage, and meniscus surfaces. **A.ii.** The MRI-based surface mesh is then reconstructed using the neural shape model, yielding a latent vector for each bone (femur, and menisci; tibia; patella) that can be used for data and statistical analyses, and a smooth surface reconstruction ideal for simulation. **A.iii**. A 24 degree-of-freedom multi-body knee model (6 tibiofemoral, 6 patellofemoral, 12 meniscal) is built automatically using the neural shape model parameterization. The knee is aligned to the reference model by registering the femur, with the tibia and patella carried by the same transform so their relative pose is preserved; ligament and tendon attachment sites (91 fibres) are updated from the reference mesh using neural shape model interpolation; cartilage and meniscus contact surfaces are automatically extracted from the smooth neural shape model surfaces; wrap surfaces are fit to bone surface regions labelled using neural shape model interpolation; and the prefemoral fat pad is created using a rule-based algorithm. **B.** The automatically built multi-body biomechanical knee model is integrated into a whole body OpenSim model which is then scaled to the gait participant using AddBiomechanics, and the Concurrent Optimization of Muscle Activations and Kinematics (COMAK) algorithm is used to simulate cartilage pressures. Gait simulations were run for 756 knees from 706 Osteoarthritis Initiative subjects through 10 different gaits each, for a total of 7,560 simulations. **C.** Data analyses were harmonized using the underlying neural shape model and its implicitly defined anatomically matching points. **C.i.** Time-varying metrics, such as mean pressure over the medial tibial cartilage, were compared between groups. **C.ii.** Neural shape models were fit to meshes over 6 years, leveraging neural shape model-based anatomically matching points to compute continuous maps of change in cartilage thickness, and aggregate statistics over groups (e.g, mean healthy, and incident OA thickness change). **C.iii.** Neural shape model-based matching points were used to determine the relationship between cartilage pressure and longitudinal change in cartilage thickness. **C.iv.** Neural shape model-based synthetic joint generation was used to explore how shape, and mechanics, systematically vary between groups.

We applied this pipeline to 756 OAI knees in three groups of 252 each (**Extended Data Table 1**): healthy knees (Kellgren-Lawrence (KL) grade <2 at baseline and four-year follow-up), incident OA knees (KL <2 at baseline, ≥2 by four-year follow-up), and mild OA knees (KL 2 at baseline, no follow-up criteria). Using longitudinal OAI data, we investigated the relationship between simulated cartilage pressures and longitudinal cartilage thinning.

**Extended Data Table 1.** Demographics and simulation convergence statistics separated by group. WOMAC = Western Ontario and McMaster Universities Osteoarthritis Index, higher WOMAC pain is more pain.

|  | Healthy | Incident OA | Mild OA |
| --- | --- | --- | --- |
| Knees (participants) | 252 (246) | 252 (233) | 252 (245) |
| Laterality (L:R) | 135:117 | 133:119 | 117:135 |
| Sex (F:M) | 136:116 | 165:87 | 141:111 |
| Age (years) | 58.6 (9.1) | 58.4 (8.3) | 60.7 (9.1) |
| BMI (kg/m<sup>2</sup>) | 27.2 (4.3) | 29.1 (4.7) | 29.5 (5.3) |
| WOMAC pain (0-20) | 1.4 (2.2) | 2.0 (3.0) | 2.1 (3.1) |
| Baseline KL (0/1/2) | 190 / 62 / 0 | 84 / 168 / 0 | 0 / 0 / 252 |
| Knees with longitudinal thickness ($\geq 2$ visits) | 247 | 251 | 244 |
| Thickness visits per knee, median (range) | 5 (2-6) | 6 (2-6) | 5 (2-6) |
| Simulations converged | 2,506 (99.4%) | 2,512 (99.7%) | 2,515 (99.8%) |
| Simulations excluded by QC | 17 (0.7%) | 21 (0.8%) | 8 (0.3%) |
| Simulations analyzed | 2,489 | 2,491 | 2,507 |

The automated pipeline (Fig. 1) successfully ran and built complete models in all 756 knees. Each knee was simulated under 10 gaits, one from a healthy adult^20^ and nine from a medial knee OA clinical trial (knee pain, KL grades 1–3)^7^, selected to span body mass, body mass index, and peak knee adduction moment. The simulations converged, yielding time-varying pressure outputs for the stance phase of gait in 99.6% of simulations (7,533/7,560), with individual gaits ranging from 99.3% to 99.9% success. Of the 7,533 converged simulations, an additional 46 were excluded as potential outliers; thus presented analyses are of 7,487 simulations.

To quantify how medial-compartment cartilage pressures differed between groups during gait, we averaged each knee’s time-varying outputs across all ten gait conditions and compared the following per-knee curves between groups: mean pressure (**Fig 2.A**), 95th percentile pressure (**Fig 2.B**), and contact area (**Fig 2.C**). Ninety-fifth percentile pressure was extracted across contacting elements at each instant, providing a measure of peak load that is insensitive to potential absolute maximum outliers. Statistical parametric mapping (SPM) showed that mean pressure was higher in healthy than in incident (0–69% of stance, p < 0.001) and mild OA knees (0–89%, p < 0.001). The 95^th^ percentile contact pressure in healthy and incident knees differed in early stance (4–21%, p = 0.006), while both exceeded mild OA around the second peak (**Fig 2.B** healthy 55-85%, p<0.001; **Fig 2.E** healthy 6.29 and incident 6.14 vs mild OA 5.94 MPa, Holm-corrected p < 0.001 and p = 0.017). Contact area of mild OA knees was significantly greater than in both other groups throughout stance (p < 0.001). Group means at each knee’s stance peaks confirmed the graded ordering of second-peak mean contact pressure (healthy > incident > mild OA, Holm-corrected p ≤ 0.008)(**Fig 2.D**). Contact area was ordered oppositely, though only mild OA differed significantly from the other two groups (both p < 0.001; **Fig 2**.F). Overall, knees with or progressing to OA had lower medial tibia contact pressure, and mild OA knees spread load over a greater surface area.

**Figure 2.**
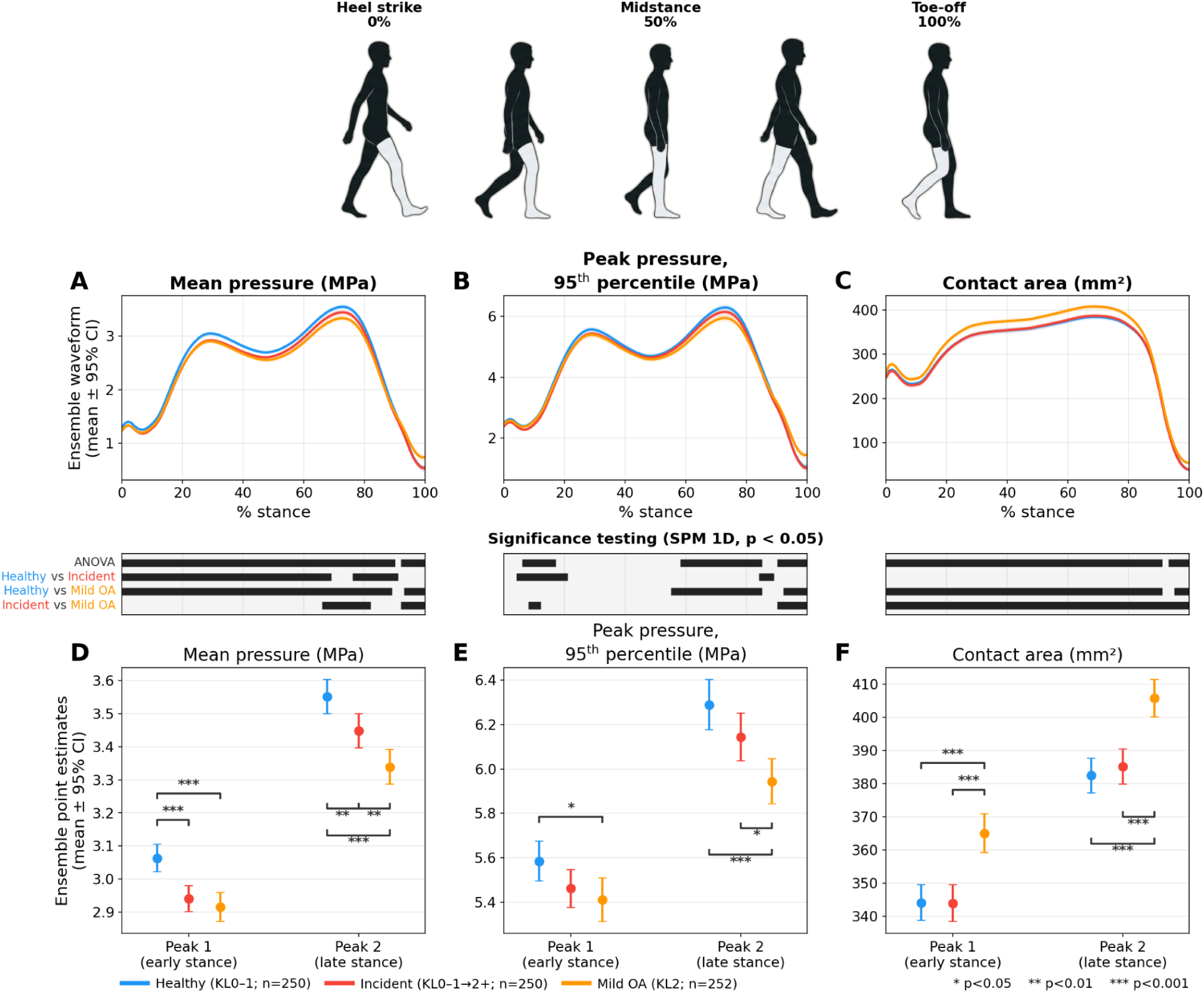
Differences amongst the three studied subgroups in medial tibial cartilage contact pressure metrics over the stance phase of gait. The diagram at the top shows the right (grey) leg throughout the stance phase of gait. **Top row**: Results from comparing the mean gait waveforms for mean tibial cartilage contact pressure (**A**), 95^th^ percentile cartilage pressure (**B**), and cartilage-to-cartilage contact area (**C**) between healthy (blue), incident (red), and mild OA (orange) groups. Gait waveforms include the mean and 95% confidence interval across knees, after each knee is first averaged over its ten conditions. Bars along the bottom of the gait waveforms show statistically different regions based on 1D statistical parametric mapping.**Bottom row**: Differences in gait metrics at the first and second peaks between groups (**D**: mean pressure, **E**: peak pressure, **F**: contact area) showing disease increases contact area and decreases pressure. Each figure includes the mean metric, 95% confidence interval, and demarcation of statistically significant differences (Holm-corrected Welch t). Analysis is of 752 knees; 250 healthy, 250 incident OA, 252 mild OA.

To understand how shape differed between groups, and how shape differences influenced cartilage pressures, we leveraged the underlying neural shape models to generate synthetic knees and then simulated their cartilage pressures during gait. We generated shapes from -3 to +3 standard deviations (SD) along the axis separating the mean healthy and the mean mild OA knees (**Figure 3**). The resulting synthetic knees showed increasing cartilage surface areas (**Fig 3.G**) from healthy (-3 SD) to mild OA (+3SD), as well as decreased medial meniscal coverage (**Fig 3.H**) and increased medial meniscal extrusion (**Fig 3.I**), both measures of meniscal degeneration. When simulating gait for synthetic knees between -3 and +3SD using the developed pipeline, the healthy knees with smaller surface area and less meniscal degeneration had lower contact area (**Fig 3.C**) and greater mean cartilage pressure throughout stance (**Fig 3.A**). Integrating generative modeling into the biomechanical pipeline enables systematic analysis of morphological effects on cartilage pressure not possible with conventional models, revealing that OA-like shape increases contact area decreasing cartilage pressure.

**Figure 3.**
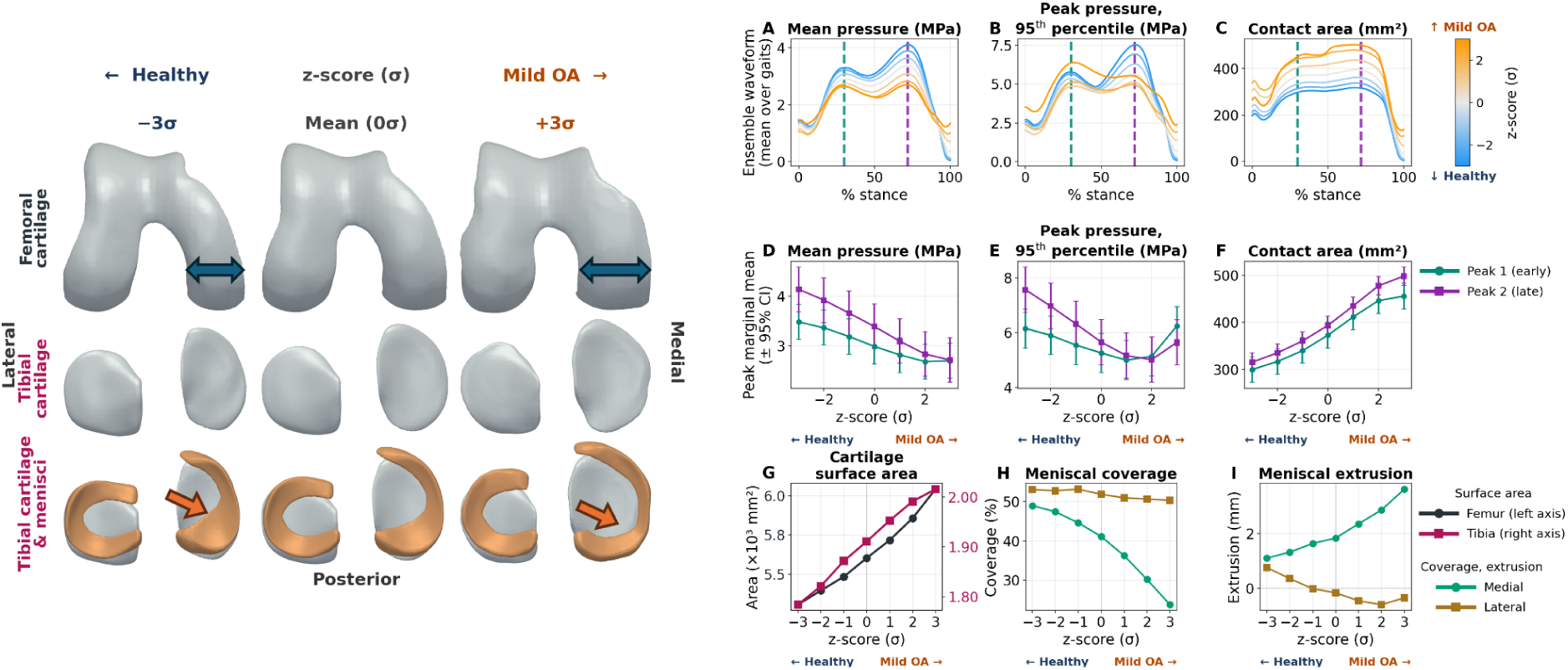
Visualization of synthetic joints along the healthy to mild OA axis and analysis of how systematically changing joint shape changes cartilage pressures. **Left:** Shows changes in femur, tibia, and meniscus shape from -3 standard deviations (healthy) to +3 standard deviations (mild OA). The more OA-like joints have large cartilage surfaces highlighted with the blue double-sided arrows, and a retracting medial meniscus highlighted with the orange arrows. **Right:** Medial tibial cartilage pressure metrics from gait simulations of the synthetic knees, averaged over the nine measured gaits. The first row plots how stance-varying cartilage pressure metrics (**A**: mean pressure, **B**: peak pressure, **C**: contact area) differ as a function of joint shape along the healthy to mild OA axis. Healthy knees (blue) show higher mean pressures, and lower contact areas, with mixed peak pressure results. Second row plots show how the same pressure metrics (**D**: mean pressure, **E**: peak pressure, **F**: contact area) for peak 1 (green) and peak 2 (purple) of stance vary as a function of shape (z-score). Quantitative analysis of the femoral and tibial cartilage surface area (**G**), meniscal coverage (**H**), and meniscal extrusion (**I**) along the healthy to mild OA shape axis. Meniscal coverage is a measure of how much of the tibial cartilage is covered in meniscus. Meniscal extrusion is a measure of how much the menisci extend beyond the tibial cartilage edge.

To determine how within-knee pressure relates to cartilage thinning, we extracted cartilage pressure metrics and 6-year cartilage thickness change for each point on the cartilage surface. Peak pressure was the maximum at each point; cumulative pressure was the time integral. Both pressure metrics were normalized to within knee deciles (0-10%, 10-20%, etc.). The relationship between normalized pressure and change in cartilage thickness (%/year) varied by cartilage region and group, with comparable results for peak and cumulative metrics (**Fig 4**). In the medial tibia, there was a graded effect between groups: the healthy group had minimal relationship between pressure and thinning (-0.07 %/year/decile, p<0.001), the mild OA group had a modest relationship (-0.14 %/year/decile, p<0.001), and the incident group had the strongest pressure thinning relationship (-0.23 %/year/decile, p<0.001) (**Fig 4.C, G**). In the medial femur, all groups had greater cartilage thinning as pressure increased, with the greatest thinning in the incident (-0.17 %/year/decile, p<0.001) and mild OA (-0.10 %/year/decile, p<0.001) groups, and limited thinning in the healthy group (-0.05 %/year/decile, p=0.002) (**Fig 4.A, E**). The lateral joint had systematic thinning differences between groups, with no meaningful relationship to pressure.

**Figure 4.**
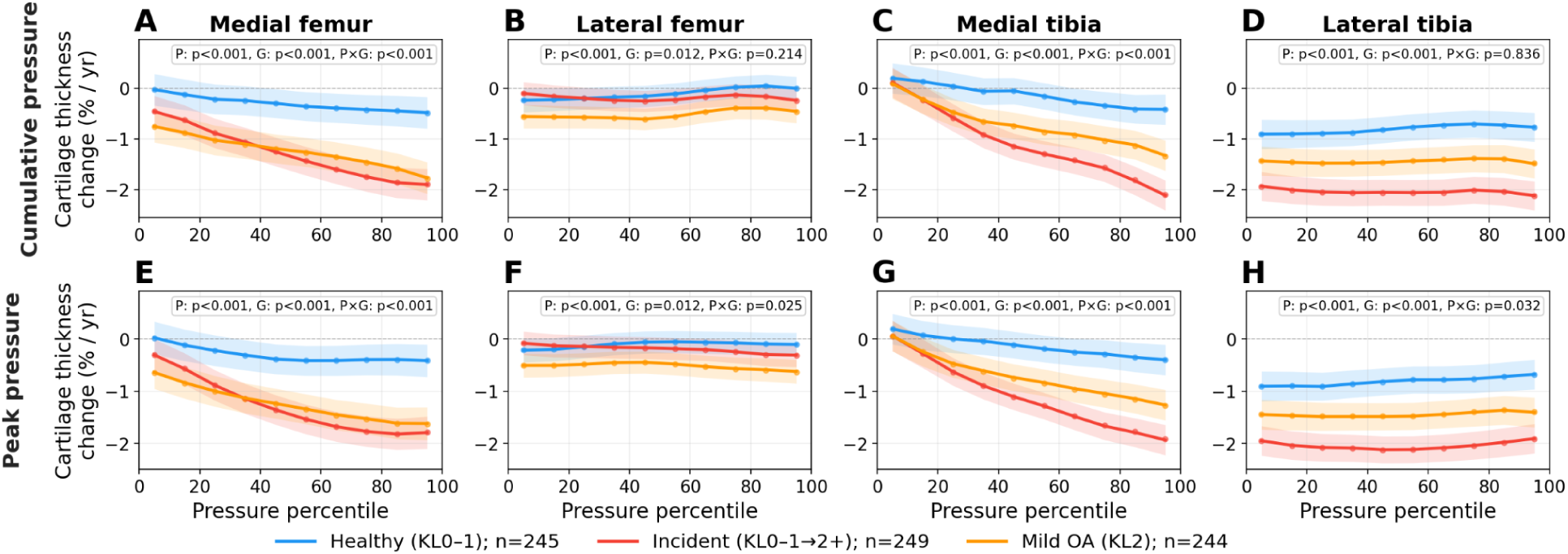
Visualization of the marginal means and 95% confidence interval for the relationship between within-knee normalized pressure (Pressure percentile) and percent cartilage thickness change per year. A negative slope indicates vertices with higher pressure had greater cartilage thinning. The top row shows results for cumulative pressure per vertex integrated over stance (MPa·s, binned in 10% increments per knee), and the bottom row shows peak pressure per vertex over stance (MPa, binned in 10% increments per knee). Significance of pressure percentile, group, and their interaction is indicated on each plot. All factors were significant (p < 0.05) except the pressure × group interaction in the lateral cumulative-pressure panels (B, D). P = percentile, G = group, PxG = Pressure x Group interaction.

To explore how between-knee spatial differences in cartilage pressure affect subsequent longitudinal cartilage thinning, we aligned simulation-based baseline pressures and individual maps of cartilage thickness change onto canonical reference cartilage surfaces using neural shape model latent interpolation (**Extended Data Figure 1**). Healthy knees had limited cartilage thinning, while incident OA knees had marked thinning in the medial femur and medial and lateral tibial plateau, and mild OA knees had thinning in these same regions (**Fig 5.E, Extended Data Figure 2**). The healthy group had greater cartilage pressure in the center of the primary contact region, whereas the incident and mild OA groups had elevated pressure in the circumferential region around it (**Fig 5.F, Extended Data Figure 2.F**). When analyzing the correlation between pressure and thinning, statistical parametric mapping showed that higher pressure in the central contact region was correlated with less thinning (**Fig 5.D and Extended Data Figure 2.D**, red), but higher pressure in the circumferential regions was correlated with more thinning (**Fig 5.D and Extended Data Figure 2.D**, blue). Pressure-based results are presented for two gaits (one healthy, one OA); pressure differences between groups and pressure-thinning correlations had similar overall results between the gaits, indicating the overarching results are robust to the exact gait analyzed.

**Figure 5.**
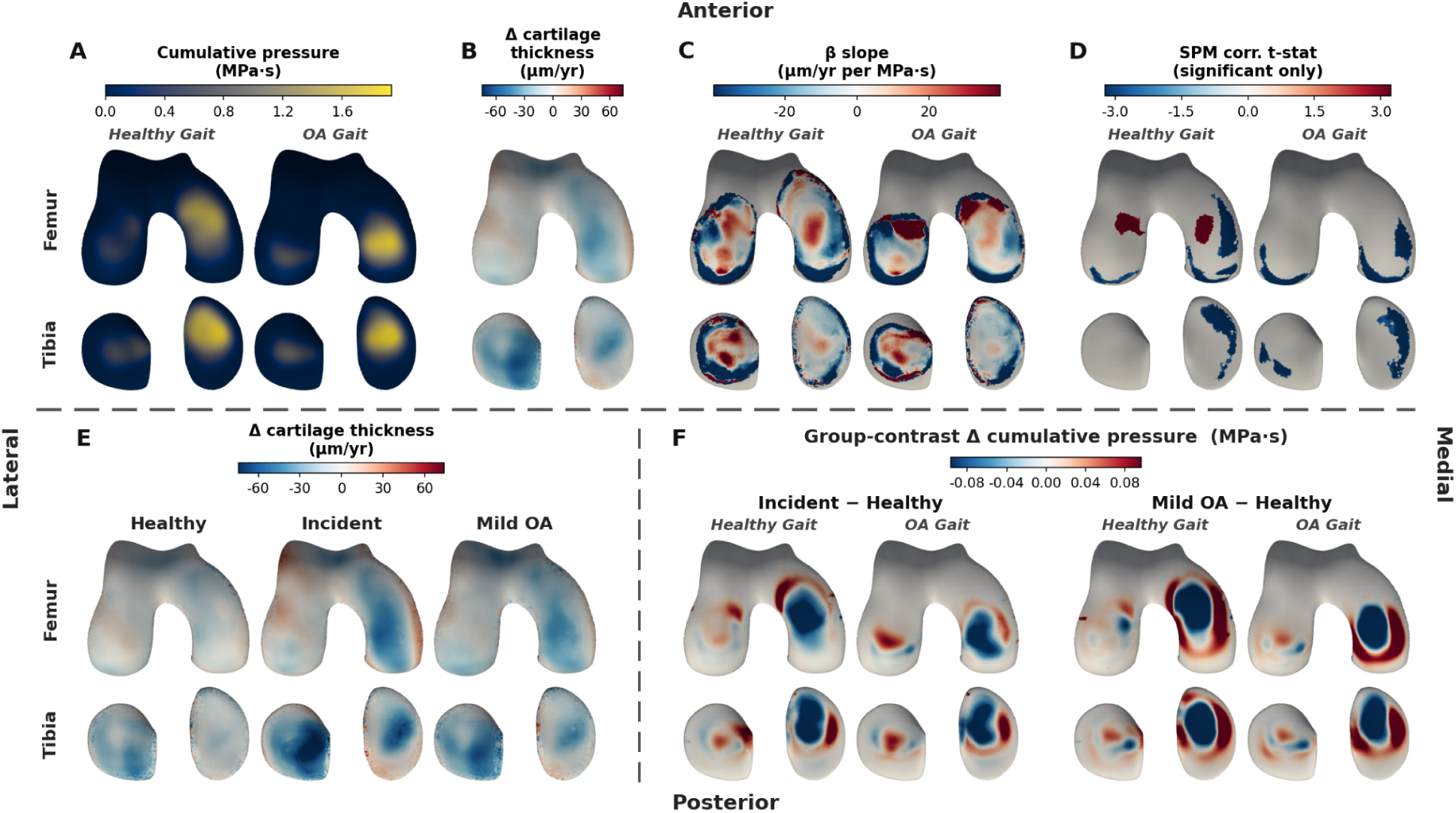
Visualization of cumulative pressure and cartilage thickness change displayed spatially over the cartilage surface. **Top. A)** Mean cumulative cartilage pressure (integrated over stance) from two representative gaits (1x healthy, 1x knee OA) where brighter yellow is higher pressure. **B)** Mean annualized change in cartilage thickness across all knees; blue is thinning and red is thickening. **C)** Point-wise slopes (β) between rate of cartilage thickness change (µm/yr) and pressure (MPa•s); blue indicates increased pressure led to more thinning, and red indicates increased pressure led to less thinning. **D)** Statistical parametric mapping showing regions where the correlation between pressure and thinning are statistically significant. **Bottom**. **E)** Change in cartilage thickness rates (µm/yr) separated by group; blue indicates cartilage thinning and red indicates thickening. **F)** Differences in pressure between the healthy group and each of the incident and mild OA groups. Blue indicates the healthy group had higher pressure, red indicates the incident/mild OA group had higher pressure. Analyses include 733 knees for the healthy gait, and 737 knees for the OA gait simulations.

The direction of association between pressure and thinning is dependent on whether it is inter-or intra-knee. Between knees, the incident and mild OA groups had lower pressure in the central contact region (**Fig 5.F**) yet greater overall thinning (**Fig 5.E**). Within a knee, the medial regions under highest pressure thinned fastest (**Fig 4**).

**Extended Data Figure 1.**
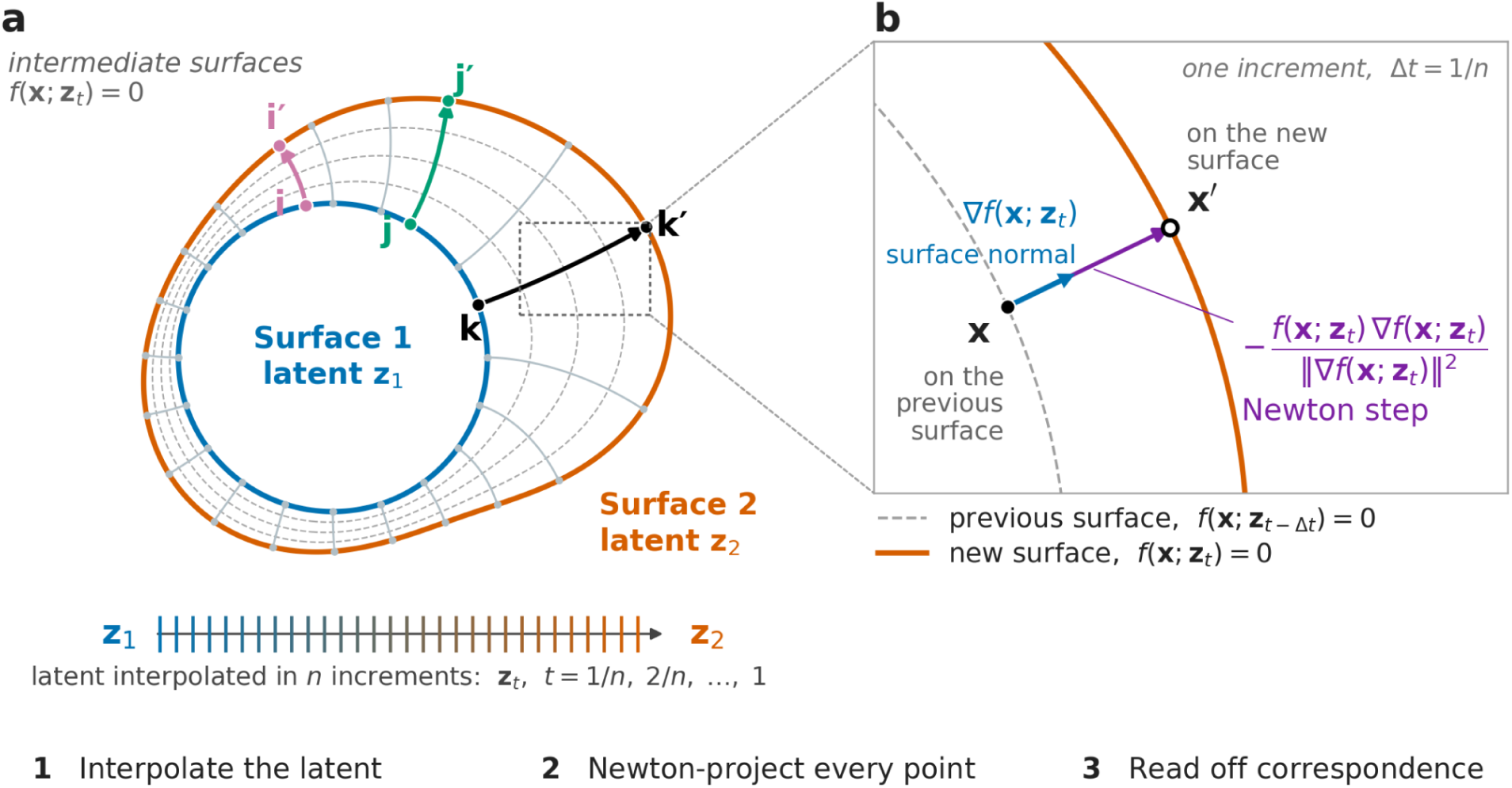
Visualization of neural shape model-based latent interpolation. The neural network f predicts the signed distance from a 3D coordinate x to the surface encoded by a latent vector z, so a surface is the level set f(x; z) = 0. a. Morphing from surface 1 (latent z₁) to surface 2 (latent z₂) over n small incremental steps (n = 100), tracking the movement of the discretized points of surface 1; each point ends at its corresponding point on surface 2 (i → i′, j → j′, k → k′). b. One step. Because f is differentiable with respect to x, ∇f(x; z_t) gives the surface normal direction and f(x; z_t)/ǁ∇f(x; z_t)ǁ gives the distance to the surface. Together they enable computing a Newton step x ← x − f∇f/ǁ∇fǁ², which is the signed distance corrected for the network’s gradient norm departing from norm=1. Evaluating f and ∇f at the same intermediate latent, and taking many small steps rather than one large one, lets each point track the deforming surface and thereby establishes point correspondence between the two surfaces.

**Extended Data Figure 2.**
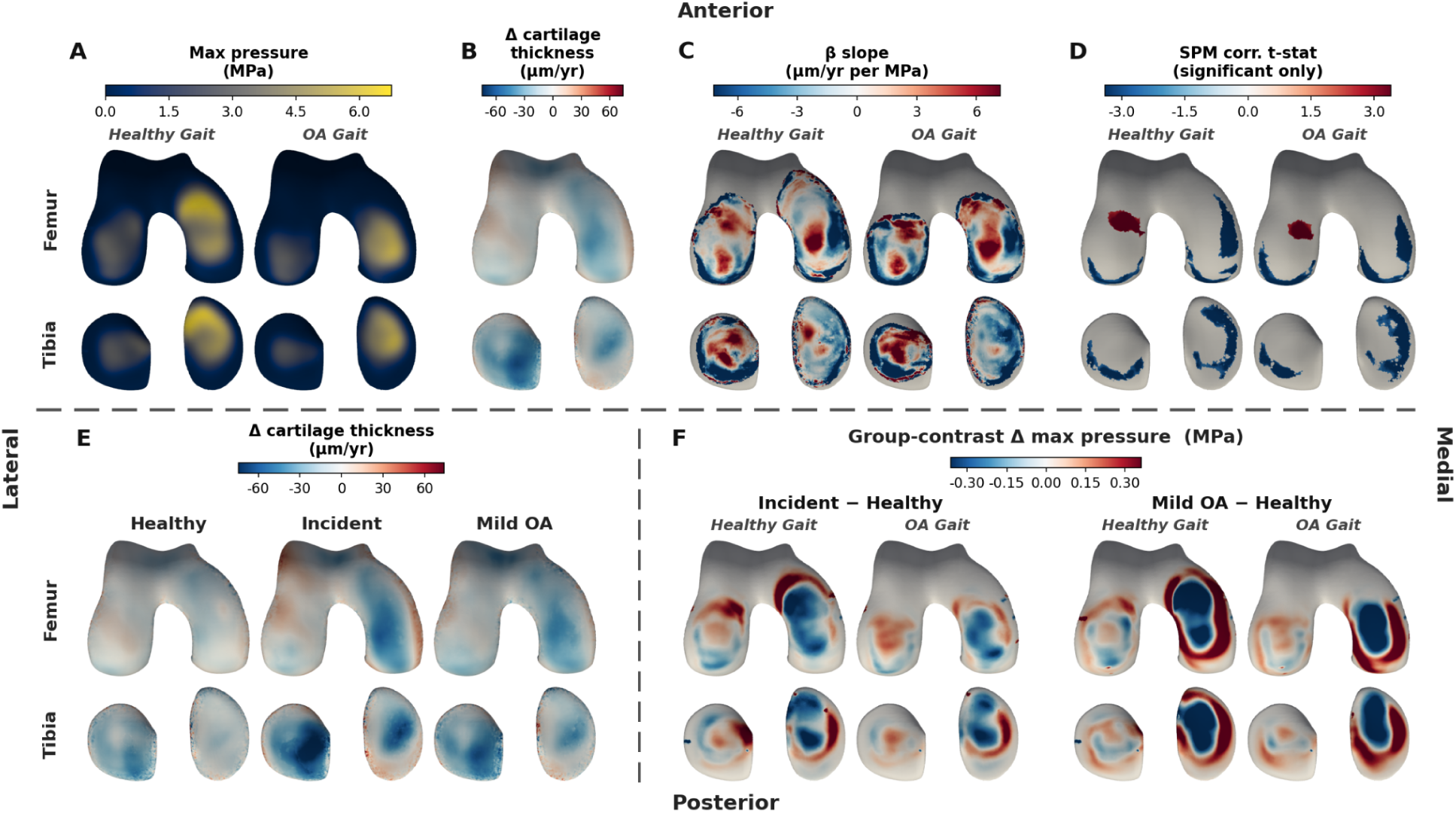
Visualization of max pressure and cartilage thickness change displayed spatially over the cartilage surface. **Top. A)** Mean max cartilage pressure (over stance) from two representative gaits (1x healthy, 1x knee OA) where brighter yellow is higher pressure. **B)** Mean annualized change in cartilage thickness across all knees; blue is thinning and red is thickening. **C)** Point-wise slopes (β) between rate of cartilage thickness change (µm/yr) and pressure (MPa); blue indicates increased pressure led to more thinning, and red indicates increased pressure led to less thinning. **D)** Statistical parametric mapping showing regions where the correlation between pressure and thinning are statistically significant. **Bottom**. **E)** Change in cartilage thickness rates (µm/yr) separated by group; blue indicates cartilage thinning and red indicates thickening. **F)** Differences in pressure between the healthy group and each of the incident and mild OA (KL2) groups. Blue indicates the healthy group had higher pressure, red indicates the incident/mild OA group had higher pressure.

Simulation results are not only dependent on joint shape but also gait differences. Correlation analyses between medial tibial cartilage pressure and gross measures of joint loading show that pressure increased with increasing knee adduction moment (**Fig 6.D, E**) and body mass (**Fig 6.F).** Visualization showed that gaits with higher external loading had higher pressure across the medial compartment and, in the lateral compartment, lower pressure centrally and anteriorly with higher pressure posteriorly (**Fig 6. G, H, I)**. While these general trends existed, within an individual gait (**Fig 6.D, E**) there was considerable variability in pressure, suggesting that joint shape-based differences have a substantial influence on localized joint loading.

**Figure 6.**
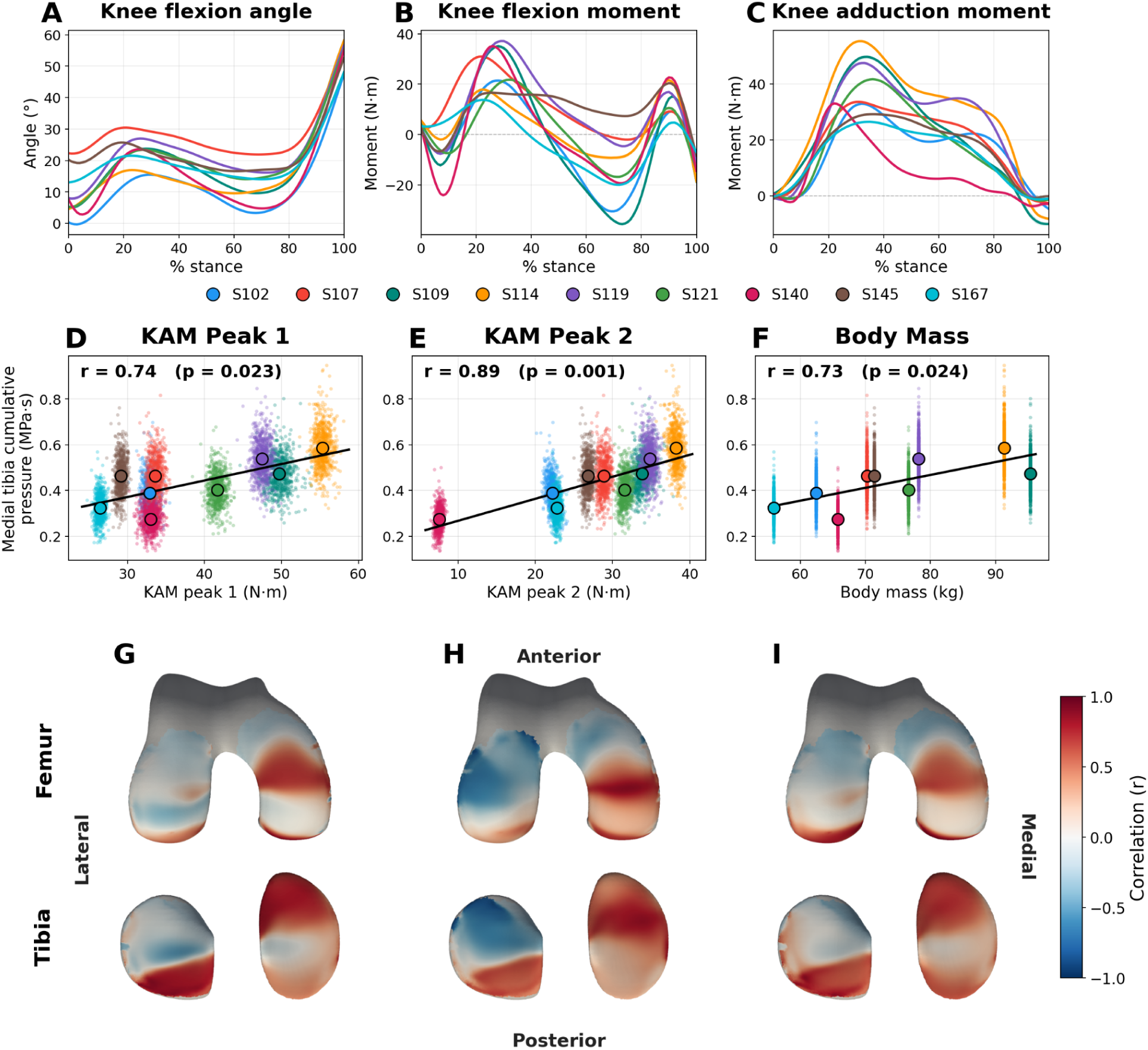
Analysis of the effects of gait versus shape on cartilage pressure. **Top.** Gait waveforms of (A) knee flexion angle, (B) knee flexion moment, and (C) knee adduction moment over the stance phase of gait for the nine knee OA clinical trial subjects. For this analysis, we use only the nine gaits from the knee OA clinical trial and exclude the single healthy reference subject to remove any effect due to study protocol. **Middle.** The correlation between cumulative pressure per cartilage vertex averaged over the medial tibial cartilage and each of KAM peak 1 (**D**), KAM peak 2 (**E**), and body mass (**F**). Each color represents a different gait. Small colored circles are individual simulation results, larger circles outlined in black are the mean pressure for that gait across all knee geometries. Small variations in KAM peaks (x-axis) are due to slight differences in simulation results with varying knee shapes; body mass has no x-axis variation as it is identical between all simulations of a single person’s gait. **Bottom**. Visualization of the spatial variations in the correlation between pressure and external loading (**G**: KAM peak 1, **H**: KAM peak 2, **I**: body mass). Red indicates a positive relationship between external loading (KAM or body mass) and pressure, while blue indicates an inverse relationship.

Cartilage pressures were related to regions of cartilage thinning (**Fig 4**), and yield between-group differences in pressure patterns (**Fig 5**). To assess whether simulated pressures can predict what knees will develop OA (incidence), we compared the ability of pressure data, neural shape model-derived joint shape, and their combination to discriminate knee OA incidence (healthy vs. incidence cohort). The best predictor of whether a knee would progress was the raw neural shape model latent vectors for the femur, tibia, and patella (AUROC = 0.69; **Figure 7**) and second best was cartilage surface shape and thickness (AUROC = 0.65; **Figure 7.A**). Pressure metrics were worst, performing only marginally better than chance (cumulative pressure AUROC: 0.56; peak pressure AUROC: 0.58). **Figure 7.B** shows that adding cumulative or peak pressure to neural shape model latents or to cartilage shape and thickness decreases predictive performance, presumably due to overfitting. Spider plots (**Figure 7.C**) reveal that both cumulative and peak pressure have variable predictive ability depending on what gait is used. Together, these data suggest that person-specific gait may enhance the ability of pressure-based metrics to predict progression.

**Figure 7.**
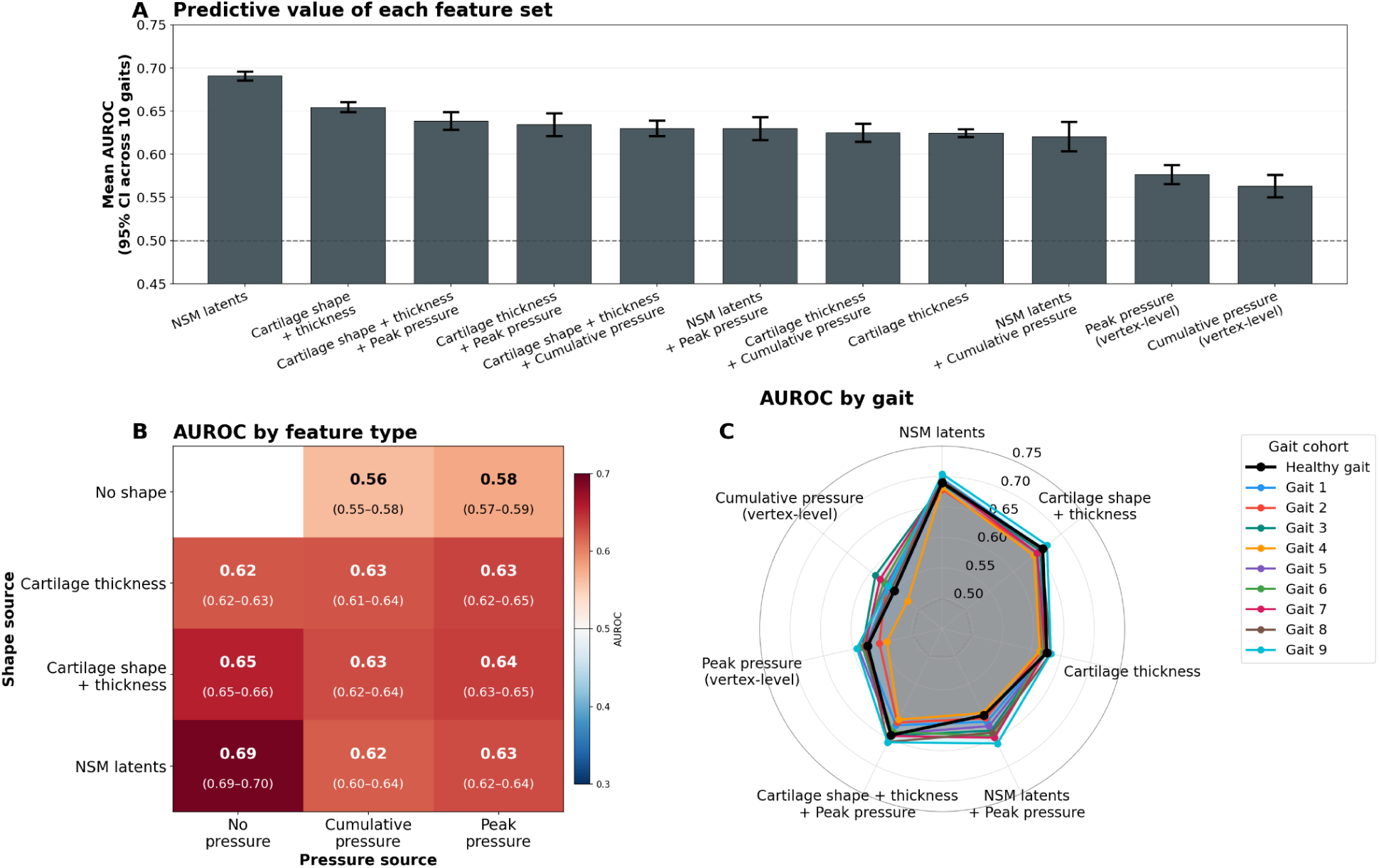
Comparison of pressure and shape-variables ability to predict whether a knee that is healthy at baseline will progress to have OA (healthy vs. incident group). **A.** The mean and 95% confidence interval for the area under the receiver operating characteristic curve (AUROC) of data derived from the 10 different walking gaits. Neural shape model (NSM) latent: neural shape model shape encoding, Cartilage shape: xyz coordinates of each cartilage vertex, Cartilage thickness: cartilage thickness at each vertex, Cumulative pressure: time-integrated pressure at each cartilage vertex, Peak pressure: per-vertex maximum over stance. **B.** Mean AUROC for combinations of predictor variables, y-axis includes individual shape-based predictors, x-axis includes individual pressure-based predictor variables. **C.** AUROC results broken down by individual gait highlighting that progression prediction varies between gaits for pressure-only metrics (cumulative pressure, peak pressure).

Cartilage thinning is conventionally assessed in an anatomically defined ‘weight-bearing’ region, rather than a mechanically defined one. In the femur, we show that using gait simulations to create person-specific weight-bearing regions improves the sensitivity to detect longitudinal cartilage thinning (**Figure 8**). The standardized response mean (SRM), a measure of how responsive a measure is to change over time, is greater for 24- and 48-month change in all three groups for the medial femur (+0.06 to +0.15 standardized response mean). The biomechanically defined region also has a greater effect size for detecting cartilage thinning in the incident and mild OA groups compared to the healthy group (+0.06 to +0.19 Cohen’s d; **Figure 8**). In the tibia, the biomechanically defined region is better able to detect between group differences in longitudinal change (+0.07 to +0.33 Cohen’s d; **Extended Data Figure 3**).

**Figure 8.**
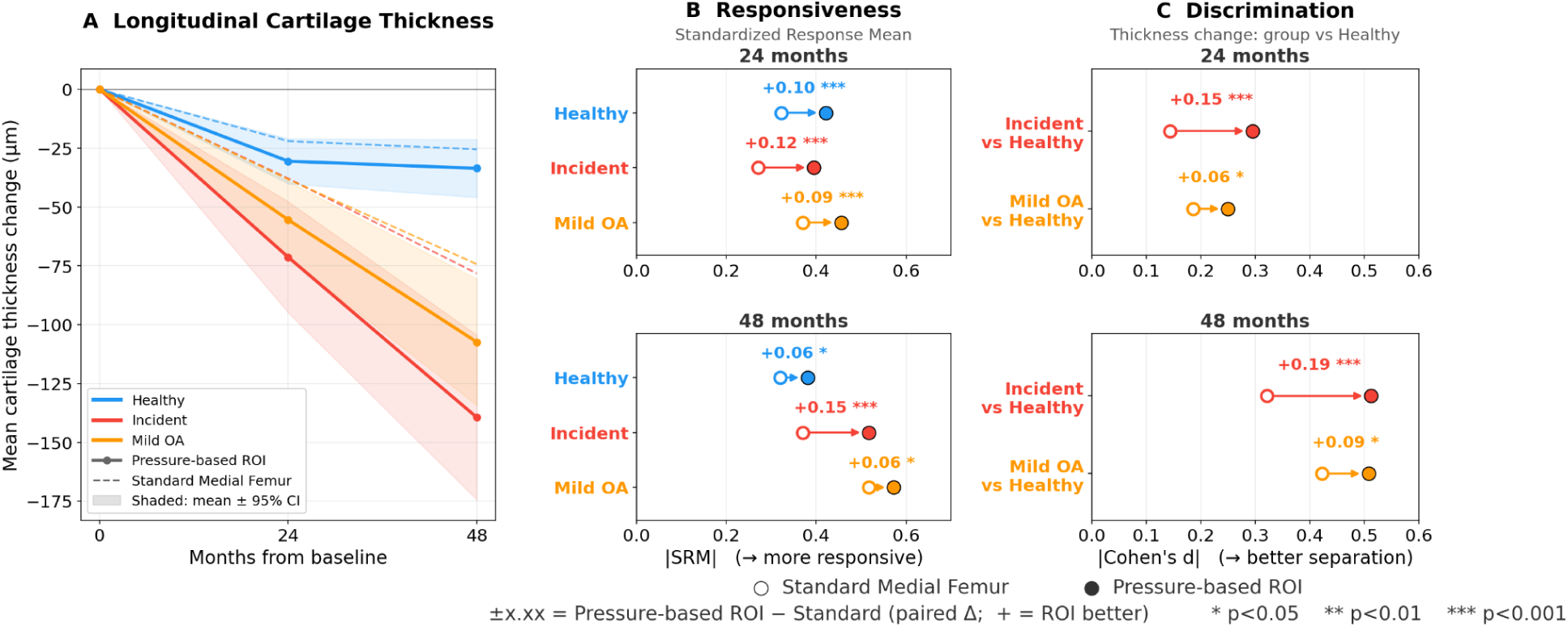
A comparison of longitudinal mean “weight-bearing” cartilage thickness changes computed using anatomically-defined vs. simulation-defined regions in the femur. **A.** Mean and 95% confidence interval for the change in cartilage thickness (µm) per group (healthy, incident, mild OA) over 48-months with simulation-defined regions (solid lines) and standard anatomically-defined regions (dashed lines). **B.** Per group responsiveness to longitudinal change for the standard (open-circle) and simulation-defined (filled circle) regions. **C.** Comparison of the ability to detect between group (healthy vs. incident, healthy vs. mild OA) longitudinal cartilage thinning using the standard (open-circle) and simulation-defined (filled circle) regions. For B and C, arrows point from the conventional to simulation-based results.

**Extended Data Figure 3.**
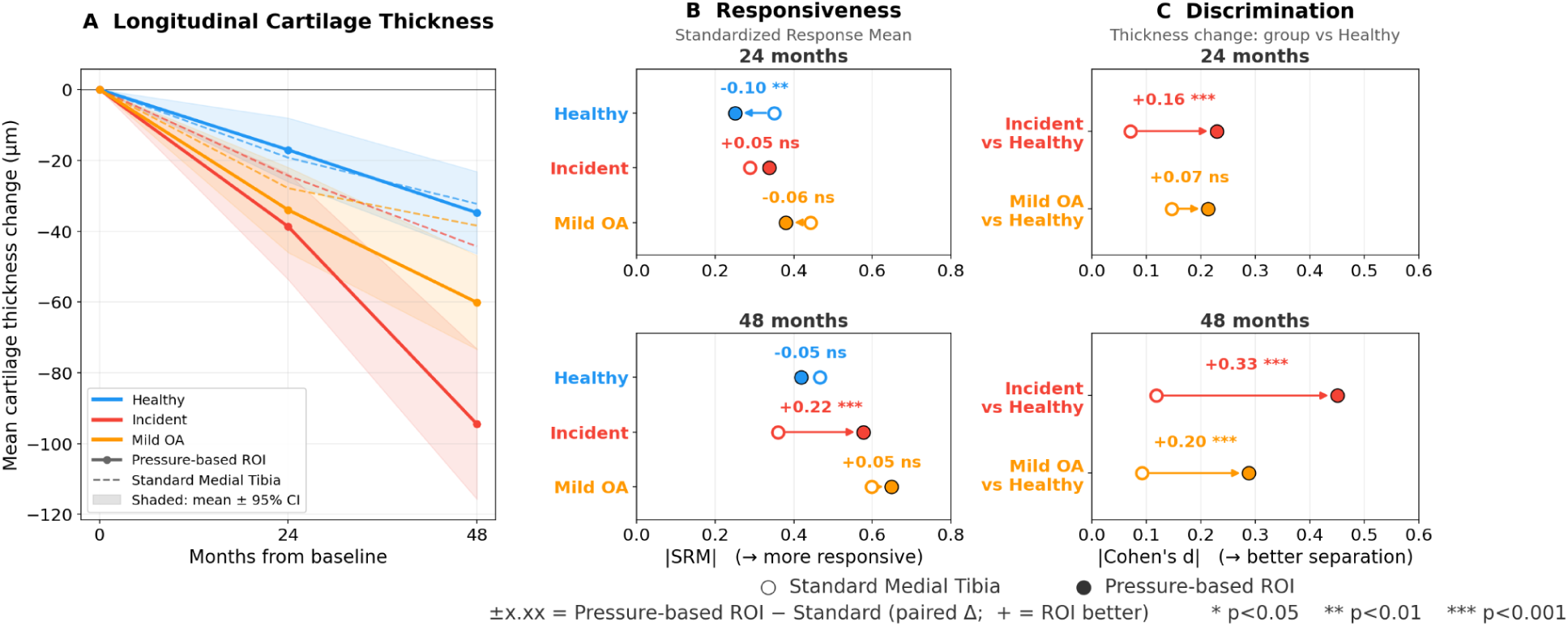
A comparison of longitudinal mean “weight-bearing” cartilage thickness changes computed using anatomically-defined vs. simulation-defined regions in the tibia. **A.** Mean and 95% confidence interval for the change in cartilage thickness (µm) per group (healthy, incident, mild OA) over 48-months with simulation-defined regions (solid lines) and standard anatomically-defined regions (dashed lines). **B.** Per group responsiveness to longitudinal change for the standard (open-circle) and simulation-defined (filled circle) regions. **C.** Comparison of the ability to detect between group (healthy vs. incident, healthy vs. mild OA) longitudinal cartilage thinning using the standard (open-circle) and simulation-defined (filled circle) regions. For B and C, arrows point from the conventional to simulation-based results.

## Discussion

Our automated neural shape model-based approach built 756 personalized knee models and ran thousands of gait simulations, providing the first large-scale analysis of how cartilage pressures relate to co-localized cartilage thickness change in knee OA. The results support the mechanical basis for knee OA progression revealing that the relationship depends on the level of comparison: within a knee, cartilage thins fastest where pressure is highest, whereas between knees, OA-related remodeling enlarges contact area and lowers pressure.

Theories of knee OA pathophysiology suggest that OA occurs and progresses due to focal changes in mechanical load beyond tissue tolerance with insufficient time for bone-cartilage remodeling^8,31^. The between-group contrasts in cartilage pressure (**Fig 5.F**) show that as OA progresses (healthy ⟶ incident ⟶ mild OA), cartilage pressures decrease in the central contact region and increase concentrically outside central contact. These changes follow from expansion of the cartilage contact area (**Fig 3**), shifting pressure onto peripheral cartilage that previously had low load (**Fig 5.F**). Importantly, SPM analysis of the pressure-thinning correlations shows that it is primarily in these regions just outside of central contact where higher pressure correlates with greater thinning (**Fig 5.D**, blue regions). Together, these data support long-held theories of knee OA <u>progression</u> where knee OA-related bone remodeling increases pressure on previously under-conditioned cartilage leaving it susceptible to cartilage deterioration and further OA progression^8^. Future work should explore how longitudinal changes in joint morphology (bone, cartilage, and meniscus shape) alter within-knee changes in cartilage tissue mechanics over time.

The finding that cartilage pressure decreased with disease severity (**Fig. 2**) is counter to the hypothesis that increased load is a risk factor for knee OA. However, given the fact that OA increases articular surface area^32,33^ it logically follows that for a fixed external gait, as in this study, increasing surface area will decrease pressure. Neural shape model-based synthetic joints in **Figure 3** show that shapes more characteristic of mild OA increase cartilage surface and contact area during gait, decreasing mean pressure, consistent with animal models of early OA^34^. Neural shape models can serve as an automated model-building tool and a generative framework for interrogating how disease or surgery change joint morphology and mechanics.

A major goal of the field is to identify treatments that slow or prevent OA. Sensitive structural biomarkers enable more efficient clinical trials (smaller sample size) or the ability to detect smaller effects. Research studies and clinical trials commonly focus on the “weight-bearing” region^35^ defined from anatomical landmarks^36,37^. In the femur, our mechanically defined region has greater responsiveness (SRM) to longitudinal thinning and yields greater between-group effect sizes than anatomy-based regions. These data suggest that personalized gait simulations may also improve the sensitivity and efficiency of knee OA clinical trials.

Alignment and body mass research suggest joint mechanics are important factors in knee OA. Varus malalignment relates to medial progression, valgus to lateral progression^38^. Recent analysis of the OAI (5,832 limbs) shows that varus alignment and body mass index (BMI) also interact to predict medial cartilage thinning, while valgus alignment and BMI independently predict lateral thinning^39^. This interaction statistically represents internal joint load (moment arm × mass). While there is a long held belief in the role of mechanics in OA, there is now evidence that fat mass alone acts on knee OA. Adipose tissue drives systemic inflammation^40–42^ and secretes adipokines implicated in cartilage degeneration^43,44^. However, systemic inflammation cannot explain compartment specificity in the body mass and alignment literature. The current finding that regions of higher cartilage pressure undergo greater thinning directly supports the role of mechanics.

Meta analyses show that higher medial knee loading during gait increases risk of OA progression, but not initiation^13^. This external loading is largely driven by body mass^45^, coronal alignment^46^, and individual motor control patterns^47^. Our data reproduce these relationships; average pressure for a given gait increases with both the knee adduction moment and body mass (**Fig 6**). However, within one gait, pressure varies between knee shapes to a degree comparable in magnitude to between-gait pressures, showing that morphology contributes to tissue-level load as much as standard external measures. Our findings mirror the meta-analysis; pressure predicts where cartilage is lost in knees that develop OA (**Fig 4**) but poorly predicts who will develop OA (**Fig 7**). Our pipeline can detect gait and shape-based effects on cartilage pressure and provides spatial information external gait metrics cannot.

Although mechanical interventions have the strongest evidence for altering OA progression, they are typically optimized using surrogates of tissue loading. Personalized gait retraining (toe-in vs. toe-out) enhances reduction of the KAM^7,48^, yet our results indicate that morphology substantially affects cartilage pressure at a given KAM. Similarly, osteotomies have shown success in slowing knee OA progression^5^, yet 2-21% of osteotomy patients go on to receive total knee replacements within 10 years^49–51^. Failures may reflect that realignment changes gross mechanics without accounting for where exactly the load is. These interventions should ideally be optimized for tissue-level pressures from each patient’s morphology and gait^52,53^. Personalized interventions could aim to reduce cartilage pressure in regions that quantitative MRI identifies as vulnerable, such as thin cartilage or cartilage with elevated T2/T1rho relaxation times indicative of degeneration. By making tissue-level pressure computable at scale, we enable studies needed to test whether optimizing against tissue-level outcomes improves results from gait retraining or osteotomy.

The primary limitation of this work is the use of multiple standardized gaits. Applying the same gaits to every knee does, however, isolate the contribution of morphology to cartilage pressure, and simulating ten gaits across all knees shows that the pressure–thinning relationships are consistent across walking patterns. The elastic foundation model provides simulated surface normal pressure but not depth-dependent tissue strain or stress. Future work could integrate FEM to include metrics such as shear stress. The elastic foundation model assumes a non-linear spring-based contact model dependent on thickness, but the current implementation uses a fixed elastic modulus. Future work could use quantitative MRI to spatially vary tissue material properties per knee. Soft tissue attachments were personalized using neural shape model correspondence; future work could segment ligament bundles or encode ligament attachments into the underlying neural shape model.

## Conclusion

Our fully automated neural shape model-based pipeline enables the largest study to date relating cartilage contact mechanics to longitudinal cartilage thinning. The pipeline automatically built all 756 knee models and successfully simulated pressures in 99.6% of 7,560 simulations. The results show that in those who progress, the regions of elevated cartilage pressures are the ones that thin the most, supporting tissue-level pressure as a local modifier of cartilage thinning. Under fixed gait, OA-related joint remodeling redistributes load, paradoxically lowering cartilage pressures. Tissue-level mechanics can now be computed for population-scale cohorts, allowing local cartilage loading to be evaluated as a determinant of disease progression.

## Methods

### Study Cohort and Disease Groups

This is a secondary analysis of data from the Osteoarthritis Initiative (OAI). The multi-center OAI study is a private-public partnership with its own institutional review board (IRB) approval and participant informed consent. We analysed 756 knees from 706 participants from the OAI. We analyzed equal sized groups of healthy, incident, and mild OA knees. Healthy is defined as not having OA (Kellgren Lawrence [KL] knee Xray grade <2) at baseline and four-year follow-up. Incidence is defined as being healthy at baseline and progressing to have OA (KL≥2) by four-year follow-up. Mild OA was defined as being KL=2 at baseline, with no follow-up criteria. We first identified all knees that fit these criteria and screened 3D renderings of their cartilage to exclude any knees with diffuse full thickness lesions as they are indicative of more severe OA than indicated by an Xray-based KL ≤ 2 reading. We used all data from the cohort with the smallest number of eligible knees (incident, n=252) and randomly sampled 252 eligible knees from the other two groups.

The demographics of the OAI knees are presented in **Extended Data Table 1**. The gait data used in this study were collected using institutional review board approved protocols with participant informed consent^7,19,20^. The healthy subject overground gait data was from a 23 year old female (1.65m, 61kg)^20^. The OA subject treadmill gait data was taken from nine subjects in a randomized clinical trial of medial knee OA^7^, which enrolled participants with symptomatic medial knee OA and KL grades 1-3 (7 female: 2 male, 68.8 years (SD = 6.7), 73.5 kg (12.8), KL1=3, KL2=2, KL3=4). The nine subjects included in this study were selected to span body mass, body mass index, and knee adduction moment peak values observed in the cohort.

### Magnetic Resonance Imaging Acquisition, Segmentation, and Surface mesh generation

We analyzed Double Echo in Steady State (DESS) knee MRIs from the OAI collected on a 3T Siemens MRI scanner with a quadrature transmit-receive knee coil (USA Instruments, Aurora, OH) with reconstructed in-plane resolution of 0.365x0.365mm, 0.7mm slice thickness, TE=5ms, TR=16ms, and flip angle=25°^54,55^. Each scan was segmented using a deep learning-based knee MRI segmentation algorithm trained on these OAI DESS knee MRI scans^23^. The algorithm has Dice similarity coefficients of 0.84-0.91 for cartilage tissues and 0.99 for bones, and average symmetric surface distance <0.25mm for all tissues. To standardize analyses, segmentations of all left knees were flipped along the medial-lateral axis.

Anatomic surface meshes were created for each of the femur, tibia, and patella bones; the femoral, medial tibial, lateral tibial, and patellar cartilage; and the medial and lateral menisci. Surface meshes were generated from the segmentation label maps using previously described methods^28^. Briefly, each tissue’s binary label map was smoothed with a Gaussian filter (variance 0.5mm for bone, 0.156mm for cartilage and menisci) and a surface was extracted with marching cubes. Each surface was made watertight and manifold and then decimated by 50%^56^. The femur and tibia long-axes were each cropped to be equal to 80% of the bone medial lateral width for shape modeling standardization.

### Neural Shape Model Surface Reconstruction and Point Correspondence

Neural shape models were fit to each bone and associated surfaces to

– extract smooth anatomic surfaces,
– interpolate keypoints from reference biomechanical models to knee-specific models,
– obtain anatomically corresponding points between knees for data analysis, and
– enable shape-modeling based interpretation

We used four neural shape models, one for the femur, tibia, and patella longitudinal modeling; each of these models included the bone and cartilage surfaces of the respective bone. These tibia and patella models were also used to build the personalized biomechanical knee models. To include the meniscus in the biomechanical model, we used an additional femur model that included the bone, cartilage, and menisci. Each model was trained on 6,239 knees from the training set of the ShapeMed-Knee dataset^28^. The neural shape model is trained to take as input a latent vector representing a specific person’s anatomy and an xyz query point and returns the signed distance of that point from the anatomic surface of interest. At the start of training, each training example is assigned a latent vector stored in a dictionary of latents. Then, during training, random points are sampled around the surface and the network weights and the latent vectors are jointly optimized to predict the signed distance of these points to each anatomic surface. A reconstruction loss minimizes the L1 norm of predicted signed distances compared to their true signed distances, while a latent regularization loss minimizes sum of squares of the latent vectors to promote a continuous multivariate Gaussian-shaped latent space.

Once trained, the neural shape model network weights are frozen, and the model is fit to a new knee’s anatomy by optimizing the latent vector to minimize the L1 norm of the network’s predicted signed distances compared to the true signed distances. The result of this optimization is a latent vector that represents the new knee’s anatomy. Neural shape model reconstructed surfaces are then generated by querying a grid of xyz coordinates with the fixed latent and running marching cubes over the returned signed distances to extract the isosurface. We used the previously described hybrid explicit-implicit network architecture^28,57,58^ that takes a global latent vector as input, applies a fully connected layer, reshapes the output to 2 x 2 x 512 and then applies five stride 2 transposed convolutions yielding three 64 x 64 plane outputs (xy, xz, yz) with 128 features each. Then, xyz point specific plane features are bi-linearly interpolated from each plane, summed, and input along with the xyz query point into a small multi-layer perceptron that outputs the requisite number of signed distances. The longitudinal model latent vectors for the femur, tibia and patella used latent vectors of length 512, the same as the original model. The femur + meniscus model used a latent vector of length 1024 to increase expressivity when encoding bone, cartilage, and meniscal shape.

### Neural shape model-based point correspondence

To transfer key points from the reference biomechanical model to a new knee, and to obtain matching anatomic points for data analysis, a latent-interpolation approach was used (**Extended Data Figure 1**). At a high level, this approach takes two latent vectors and iteratively flows the vertices from the surface encoded with the first latent vector onto the surface encoded by the second latent vector. This is possible because the neural shape model (f) is a differentiable function that maps a query point (x, y, z) and a latent vector to the signed distance from that point to the latent encoded surface, so its spatial gradient ∇f gives the surface normal. The latent vector of the first surface is advanced to the latent vector of the second surface via interpolation using 100 steps. At each interpolation step, every point is projected onto the surface of the intermediate latent with a single Newton step, x ← x − f(x)·∇f(x) / ǁ∇f(x)ǁ², which moves the point along the surface normal by f(x)/ǁ∇f(x)ǁ. Because both the step direction (∇f) and the step size (f) are evaluated at the same intermediate latent, and the latent changes only slightly per increment, the points track the deforming surface; the endpoint of this flow is the corresponding point on the second surface.

### Personalized Biomechanical Knee Model

We built personalized biomechanical knee models based on a previously developed and validated OpenSim-based musculoskeletal model with extended soft tissue contact using an elastic foundation model^19,20^. The musculoskeletal model includes a 24 degree of freedom knee: 6 tibiofemoral, 6 patellofemoral, 6 medial tibio-meniscal, 6 lateral tibio-meniscal. The knee includes 91 nonlinear elastic fiber elements modeled using the Blankevoort nonlinear springs from OpenSim as described previously^59^. We use the ligament specific linear stiffness, toe region, and damping coefficients from the reference model^20^, with the exception of the patellar tendon, which uses a higher linear stiffness of 12,000 N per fiber in line with previous knee models^15^ and experimental studies of patellar tendon tissue properties^60–62^. Ligament attachments were determined using the neural shape model-based point correspondence approach. Each neural shape model was first fit to the reference validated model’s anatomic surfaces to obtain a latent vector and each ligament attachment xyz coordinate was documented. Then, for each new knee to be modeled, these attachment points were interpolated from their xyz surface coordinate associated with the reference latent to the surface associated with the new knees latent.

**Cartilage-cartilage and cartilage-meniscus contact** used the same contact pairs and tissue properties as the reference model^20^. Contact used an elastic foundation model^63^ where contact is detected at each cell of the tissue surface by projecting rays in the negative of the surface normal to detect penetration. When penetration is detected, the cartilage-cartilage contact pressure uses a variable-property formulation which uses separate elastic foundations for each surface that are dependent on local thickness, elastic modulus (E), and Poisson’s ratio (v).

Cartilage assumes E= 5MPa and v=0.45, with spatially varying knee-specific thickness from the MRI-derived cartilage and bone surfaces. Cartilage-meniscus contact used a lumped-parameter elastic foundation model (averaged modulus/Poisson’s ratio, summed layer thickness) with a linear pressure-depth law, E = 5MPa, v=0.45, and 3mm meniscal thickness.

#### Contact surface extraction

To enable appropriate contact modeling, we extract the articular surfaces of the femoral, tibial, and patellar cartilage, and a superior and inferior surface of each menisci. To extract the cartilage surfaces, first, each neural shape model reconstructed surface was resampled to have uniform density of 3 triangles/mm^2^ ^16^. Then vectors were projected in the negative of the normal direction for each point on the cartilage surface and if the vector intersected the underlying bone it was kept, all other vertices were removed. To remove aberrant triangles, the single largest connected component was retained, boundary cells with only one edge-neighbor were removed, and the surface was Laplacian smoothed (100 iterations).

Menisci-cartilage contact requires extraction of the superior surface of the meniscus for femoral-meniscal contact, and the inferior surface for tibial-meniscal contact. The meniscal surface is first built the same as in the cartilage, except it was resampled to have a uniform density of 4 triangles/mm^2^. Candidate vertices are identified by projecting vectors normal to the meniscal surface, and if they intersect the femur bone then they are retained for the superior surface and if they intersect the tibia bone they are retained for the inferior surface. The same largest connected component, removal of isolated boundary cells, and Laplacian smoothing was performed as in the cartilage, except smoothing included 10 iterations. The outer vertical border of the menisci (lateral side of lateral meniscus, medial side of medial meniscus) can inadvertently be retained and cause large unintended contact forces. To remove these points, the menisci vertices are expressed in polar coordinates and the 95^th^ percentile radius of all points in 100 angular bins is computed and filtered. Points outside of this 95^th^ percentile are removed and the largest connected component is again extracted and isolated cells removed.

#### Prefemoral fatpad

The reference model trochlear cartilage extends superiorly beyond the physiologic boundaries of the trochlea. This provides a contact surface for the patellar cartilage in terminal knee extension, preventing dislocation. Instead of creating this artificial cartilage surface, we generate a new contact surface that represents the prefemoral fatpad, the physiologic tissue that the patella would contact if it translated superior to the trochlea (**Extended Data Figure 4**). The fatpad was created by extending the exposed anterior femoral bone surface (regions not covered by cartilage) outward along its surface normals by 1–6 mm (1mm at the top of the trochlear cartilage, rising logarithmically to 6mm at the most superior point), with the inflation at each vertex limited by ray-cast clearance to the patella so the surface approached but did not penetrate the patella. The resulting surface was cropped to the region within 25 mm of the patella, smoothed, and resampled. The fat pad was assigned an elastic modulus of 4 MPa, a Poisson’s ratio of 0.45, and a constant thickness of 10 mm, and was modeled as a nonlinear elastic-foundation (lumped) contact between the femur-attached fat-pad surface and the patellar cartilage. This compliant anterior contact cushions the patellofemoral joint to prevent non-physiological penetration of the patella into the anterior femur during simulation.

**Extended Data Figure 4.**
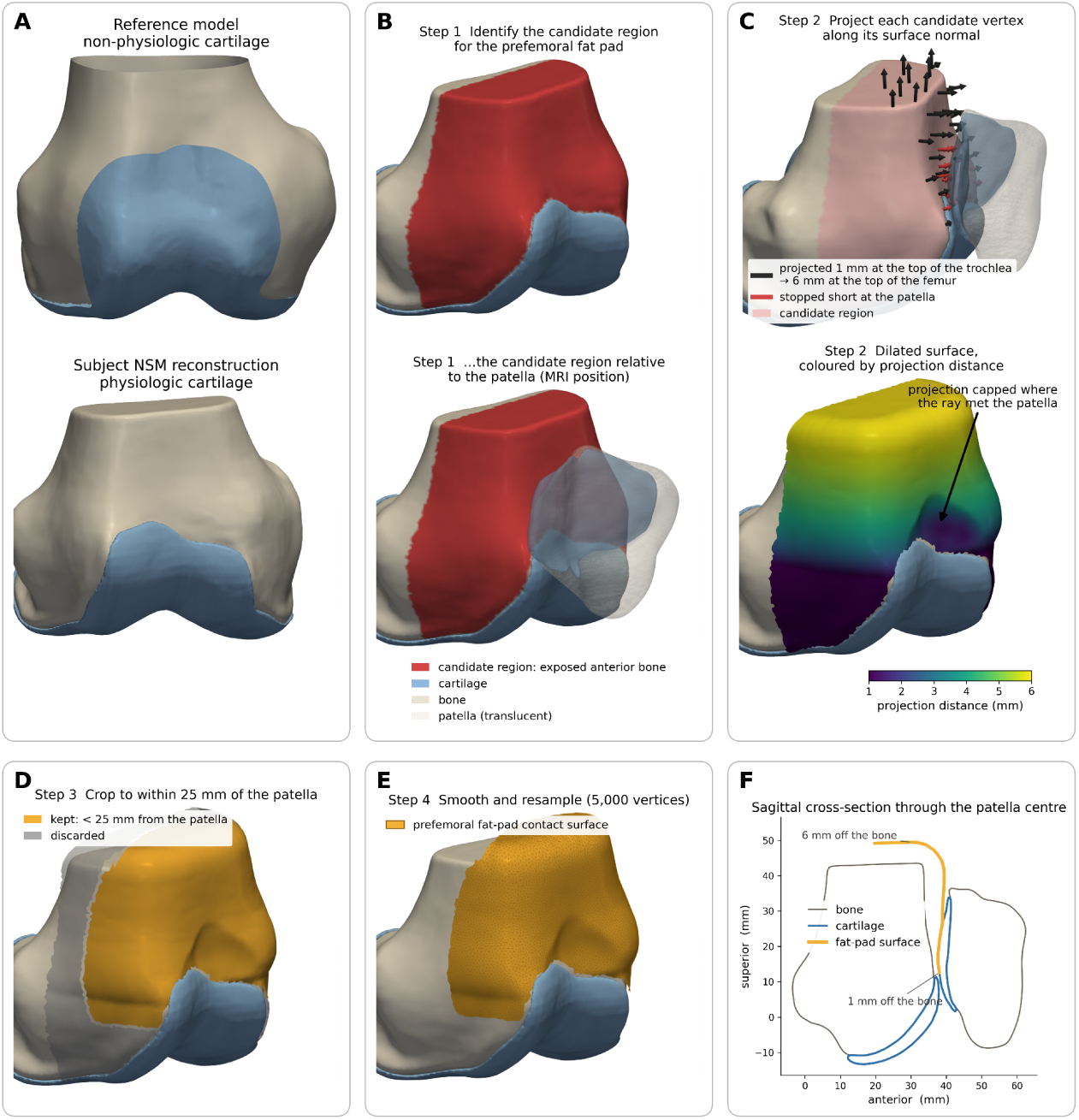
Construction of the prefemoral fat-pad contact surface. A. The reference model’s trochlear cartilage (top) is extended superiorly beyond the anatomic trochlea so that the patella has something to contact in terminal extension; a knee’s neural-shape-model cartilage (bottom) ends at the trochlea. B. The candidate region for the fat pad: vertices of the femoral bone surface not covered by cartilage and lying in the anterior half of the bone (red), shown without (top) and with (bottom) the patella in its MRI position. C. Top: each candidate vertex is projected along its surface normal, by 1 mm at the top of the trochlea increasing to 6 mm at the top of the femur; where a ray cast along the normal meets the patella the projection is stopped short so the surface does not pass through the patella (red). Bottom: the resulting dilated surface coloured by projection distance. D. The dilated surface is cropped to vertices within 25 mm of the patella. E. The cropped surface is smoothed and resampled to 5,000 vertices, giving the fat-pad contact surface. F. Sagittal cross-section through the centre of the patella showing the fat-pad surface standing 1–6 mm off the exposed anterior femur, between the bone and the patellar cartilage.

**Wrap surfaces** were personalized for each knee using an optimization-based approach. On the reference bones, each vertex was labeled relative to every reference model wrap surface as inside or outside the wrap, and as near-surface if within 0.5mm (femur, tibia) or 1mm (patella). These points, and their labels, were then transferred to each knee’s neural shape model reconstructed bone using neural shape model interpolation. Each wrap primitive (ellipsoid, cylinder) was then fit to the knee’s labelled points by optimizing its parameters so that vertices inside of the reference wrap remained inside and those outside remained outside using a squared-hinge margin penalty on the fitted wrap’s signed-distance field, while a regularization term kept each wrap near its reference pose and size. Wrap parameters were optimized using the limited-memory Broyden–Fletcher–Goldfarb–Shanno (L-BFGS) algorithm, preceded by a brief Adam warm-up for ellipsoidal wraps.

#### Model Scaling

All of the above steps were performed after the knee’s femur bone was similarity registered (rigid plus isotropic scale) to the reference model and the fitted transformation was applied to the tibia, patella, and menisci. This removed pose and size differences, yielding models that differed only in shape and inter-bone alignment. To simulate gait for a subject other than the reference, the model’s knee was reduced to a hinge, its arms and OpenSim Joint and Articular Mechanics (JAM) components were removed, and this skeleton was scaled to the gait participant’s motion-capture markers and ground reaction forces with AddBiomechanics^64^, yielding per-body scale factors and inertial parameters. These factors were applied to the reference-size model using standard OpenSim scaling: each body was scaled by its AddBiomechanics per-axis factors, while the knee bodies (femur, tibia, patella, and menisci) and their ligament/tendon attachments and wrap surfaces were scaled isotropically by the mean of the femur and tibia long-axis scale factors. After scaling, all ligament slack lengths were updated to maintain the same strain as the reference model.

The knee model was then finalized by running a settling and a sweep simulation. Before these, each simulation checked patellar stability using a custom forward simulation; if the patella excessively translated or the simulation failed to converge, the reference patella position was translated 4mm inferiorly and the check repeated; 60 of 7,533 completed simulations translated the patella (53 by 4mm, 7 by 8mm). In the settling simulation, a forward simulation was run with every muscle’s force set to a constant 1% of its maximum isometric force and all degrees of freedom locked except the secondary knee kinematics, allowing these coordinates to settle to equilibrium. This was repeated twice, each time updating the ligament slack lengths so that each ligament reached its reference strain at the settled pose. A forward simulation then flexed the knee from 0° to 60° over 0.6 s under the same 1% muscle forces. Spline functions relating each secondary coordinate to knee flexion were fit to this sweep and added to the model, producing an interim (“constrained”) model in which the secondary knee kinematics are prescribed functions of knee flexion. This interim model was used for traditional inverse kinematics of each subject’s gait, after which the secondary degrees of freedom are released and solved concurrently with muscle forces and contact in the full simulation.

#### Synthetic Joint Generation

Synthetic joints are defined by a per-knee 2069-dimensional joint descriptor vector, the same parameterization computed for each of the 756 knees in the cohort. The vector is comprised of a 1024 femur and meniscus shape vector, 512 tibia shape vector, 512 patella shape vector, and a 7-dimensional pose vector per body including the Euler rotations (3), translations (3) and a scale factor. Pose features are defined relative to the population mean size and position. To create a new joint, the vector is broken into its components. Each shape vector is decoded by its respective neural shape model to reconstruct anatomic surfaces. The pose components are recomposed into a similarity transform per body that positions, orients, and scales each reconstruction, with the femur transform defined in absolute terms and the tibia and patella transforms defined relative to the femur, so the reconstructions form a physiologically posed joint rather than three independent bodies. Then, the standard model building steps are performed, including interpolating ligament and tendon attachments and wrap surfaces, building the prefemoral fat pad, and extracting the cartilage and meniscal contact surfaces and resampling them.

### Gait Simulations with Cartilage Pressure

We used the Concurrent Optimization of Muscle Activations and Kinematics (COMAK) algorithm to simulate gait for each personalized knee geometry, obtaining shape and gait-specific internal knee contact pressures, muscle activations, and secondary knee kinematics. We ran the COMAK pipeline for all combinations of 10 gaits (1 healthy, 9 OA) and 756 knee geometries. Simulations were run for the stance phase of gait. To obtain kinematics for the COMAK algorithm to track, we first ran inverse kinematics using 1 degree of freedom interim knee model from the sweep simulation. Then, we ran the full COMAK algorithm, which, at each time step, concurrently optimizes the muscle activations and secondary knee kinematics (the tibiofemoral translations and secondary rotations plus the patellofemoral and meniscal degrees of freedom) required to reproduce the measured primary joint kinematics and net joint moments while minimizing muscle effort, with cartilage contact and ligament forces resolved as part of the solution. Specifically, COMAK minimized the sum of the muscle volume normalized muscle activations squared with per muscle weightings, where a weight >1 penalized use of that muscle and a weight <1 encouraged use of that muscle. We used muscle weights as previously described^20^. The nominal weighting was 1, except for: lateral gastrocnemius=7, medial gastrocnemius=4, rectus femoris=3, biceps femoris long head=2, biceps femoris short head=2, semitendinosus=2, semimembranosus=2, soleus=0.9, gluteus medius=0.9, gluteus minimus=0.9. These weightings are aligned with other work that targeted matching simulated joint reaction forces with measured joint reaction forces in individuals with instrumented total knee replacements^65,66^.

When using fully personalized geometries on 20 test knees, simulation times took a median of 5 hours for 13 test knees, with 7 knees not completing within time cutoffs. To improve convergence and accelerate simulations, we changed convergence criteria to be generalized force units computed by multiplying accelerations by the mass/inertial parameters of each degree of freedom and setting the optimization threshold to 1 unit (N or Nm). We also increased meniscal moments of inertia from 10^-5^ → 10^-4^ kg·m^2^. The default simulations searched for contact between surfaces at a distance of 20mm. We reduced search distances to be contact surface specific based on empirical simulation data: tibiofemoral=6mm, patellofemoral=5mm, meniscal=2mm, fatpad=8mm. This both reduced the search space and limited spurious, unphysiologically deep contact penetrations. COMAK max outer iterations were set to 25 and ipopt max iterations were set to 500. Using this updated pipeline, all 20 test simulations completed in a mean=33 mins, standard deviation=8.3min (minimum=19.2minutes, maximum=53.4minutes). Compared to the 13 simulations that completed with the default settings, the median per trial root mean squared error was: 0.10-0.78° for knee rotations, 0.09-0.15mm for knee translations, 6.0x10^4^ Pa for contact pressure (0.86% of peak contact pressure of ∼7x10^6^ Pa), 6.3 N for muscle forces, 0.009 for muscle activations (<1% of 0-1 range), and 0.97 Nm for joint moments.

### Cartilage Thickness, Cartilage Pressure, and Spatial Coregistration

The neural shape model was used to co-register all cartilage thickness and pressure data. To enable this, in addition to fitting the neural shape model to each knee’s baseline bone/cartilage surfaces for simulation purposes, we fit the neural shape model to anatomic surfaces for every subsequent OAI visit for which MRI data was available. The same segmentation and surface extraction methods were used for those data. For thickness analysis, we reconstructed the knee’s bone and cartilage surface using the neural shape model, and then computed thickness on the cartilage articular surface by projecting vectors in the negative of the normal direction and calculating the Euclidean distance to the underlying bone if the ray penetrated it, otherwise assigning a thickness of 0mm. Cartilage thickness data were available for a median of 5 timepoints/knee (mean 5.0). Fourteen knees (1.9%) had thickness at only one visit and were excluded from thinning analyses. The number of knees with 2-6 timepoints were: 2 = 38 (5.0%), 3 = 55 (7.3%), 4 = 103 (13.6%), 5 = 193 (25.5%), 6 = 353 (46.7%).

Baseline cartilage pressures, and cartilage thickness from every timepoint were mapped onto a common 20,000-vertex reference cartilage surface for each bone using neural shape model latent interpolation. For each knee, the reference and personalized surfaces were placed in dense correspondence by interpolating between the population-mean (zero) latent and the personalized knee’s latent, and the per-vertex scalar (thickness or pressure) was transferred by a distance-weighted average of its three nearest neighbors.

Once all thickness data were aligned in the canonical representation, change in cartilage thickness was computed as the slope of the ordinary least squares fit of thickness (mm) vs. time (years) computed independently for each vertex and knee. Normalized change in cartilage thickness was computed as the % of the knee and vertex specific baseline thickness.

Mean pressure and mean thickness were computed over regions of interest. For tibial cartilage, the medial and lateral values represent the whole cartilage surface of that respective region. For femoral cartilage, medial and lateral metrics were computed over the condyles only, excluding the trochlea. The condyles were considered any point posterior to the landmark where trochlear cartilage separates into the two condyles at the intercondylar notch^37^.

### Data and Statistical Analysis

Contact pressure data were analyzed at each timepoint, over stance, and at each vertex within the contact area. Timepoint metrics were computed by analyzing the contacting elements of a region (e.g., medial tibia) at a specific timepoint along the normalized stance curve and include the mean and peak (95th percentile) pressure (Pa). Vertex level metrics were computed by analyzing all timepoints (over stance) for a specific vertex and include the max pressure (Pa) over time as well as the integral of pressure over time (Pa·s).

Outliers in pressure metrics were considered simulation failures and were excluded. A total of 36 simulations were identified as vertex-level outliers, and 31 simulations as time-series level outliers. These two groups had large overlap, yielding a total of 46 simulations (0.61%) of 7,533 as outliers, including 17 healthy, 21 incident, and 8 mild OA simulations.

#### Between Group Comparisons in Cartilage Pressure

We used one-dimensional statistical parametric mapping to assess where pressure and contact area differed between the three groups (healthy, incident, mild OA) over the stance phase of gait. For each OAI knee, we first computed an ensemble of all 10 gaits as the unit of analysis. We used a three-group omnibus one-way analysis of variance to determine if there were group differences, then pair-wise two sample t-tests to assess which groups differed. Both analyses were done with alpha=0.05 for cluster-extent inference using random field theory^67^. To analyze differences in peak pressure, we extracted the time of the maximum of the mean pressure for early stance (0-50%) and late stance (50-100%) and extracted mean pressure, peak pressure, and contact area at this timepoint. Pairwise differences were computed using Welch’s t-test with a Holm-correction for multiple comparisons.

#### Synthetic Joints

Synthetic joints along the healthy to mild OA axis were created using the 2069-dimensional joint descriptor vectors of each knee. The healthy-to-mild OA shape axis was defined as the unit vector connecting the mean of the mild OA joint descriptors and the mean of the healthy joint descriptors. All knee’s joint descriptors were then projected onto this vector and the mean and standard deviation was computed. Joint descriptors were then extracted at -3 to +3 standard deviations and were used to generate synthetic knee joints used for data and statistical analysis. Synthetic knees were simulated using only the nine gaits from the knee OA clinical trial, excluding the healthy reference gait to remove systematic differences between studies.

The same as for the between group pressure analysis, we extracted the time of the maximum of the mean pressure for early stance (0-49%) and late stance (50-100%) and extracted mean pressure, peak pressure, and contact area. These were modeled as dependent variables using a linear mixed effects model with a fixed effect of sigma as a categorical variable and random effect of gait. Marginal means and their 95% confidence intervals were then plotted as a function of sigma. A second version of the model with sigma as a continuous variable was used to estimate the relationship between sigma and pressure/contact area. Mixed effects models used restricted maximum likelihood (REML) from the lme4 package.

#### Within Knee Cartilage Pressure and Cartilage Thickness Change Relationships

We modeled the relationship between cartilage pressure and cartilage thickness change using one mixed effects model per region (medial tibia, lateral tibia, medial femur, lateral femur) and pressure metric (cumulative pressure, peak pressure). Analyses included vertices with pressure > 0 Pa. To improve signal to noise ratio, vertices were binned into deciles (0-10%, 10-20%… 90-100%) based on the pressure metric of interest within each knee, gait, and region; results from all 10 gaits were included in the same model. The dependent variable was the area weighted mean cartilage thickness change (%/year) for the vertices in a given bin, fixed effects included pressure decile, group (healthy, incident, mild OA), and their interaction; decile was modeled categorically to enable modeling non-linear relationships between pressure and thinning. Random effects included knee intercept and slope, gait intercept, and a knee × pressure-decile random intercept. Visualizations included marginal means per decile and group and their 95% confidence intervals. Secondary analysis used a mixed effects model but treated decile continuously to estimate the relationship between pressure and thinning, and used random effects of knee intercept and slope, and gait slope. Fitting used restricted maximum likelihood in lme4.

#### Statistical Parametric Mapping

Continuous spatial analysis of the relationship between pressure (cumulative, peak) and cartilage thickness change (mm/year) was performed separately for the femur and tibia using permutation-based statistical parametric mapping. Only vertices with data from >50% of knees were included in the analysis. Correlations were computed per vertex, converted into t-statistic maps, and a cluster forming threshold was set to |t| = 2.33 (each tail p<0.01). Cluster significance was computed for cluster surface area (summed cell areas). The cluster-level null distribution was determined using 10,000 permutations and cluster alpha was set to 0.05, or the 95^th^ percentile of the max-cluster-area distribution from permutations. This same analysis was run for two gaits, the healthy reference subject, and one subject with KL grade 3 knee OA.

#### Loading Determinants of Pressure

To assess how changes in gait affected cartilage pressures, for each gait we extracted the KAM peak 1 as the max over the first 50% of stance, and KAM peak 2 as the max over the last 50% of stance. For each gait, the body mass of the subject, and the mean KAM (peak 1, peak 2) and mean pressure metric across all included OAI knee simulations were extracted for analysis. Analysis included the nine gaits from the same knee OA cohort to remove systematic differences between studies. The primary analysis was the simple Pearson correlation between each of KAM peak 1, KAM peak 2, and body mass with the mean cumulative cartilage pressure. Visualizations of the continuous spatial relationship between external loading and pressure were generated by computing the per-vertex Pearson correlation between the nine gait cumulative pressures and the respective covariate. Visualizations only include vertices where >50% of the cohort’s knees have data.

#### Predicting Knee OA Progression

To assess whether computed cartilage pressure metrics can predict who will progress, we performed logistic regression analyses to discriminate between the healthy and incident cohorts. Logistic regression inputs were comprised of a shape component (None, per vertex cartilage thickness, per vertex cartilage thickness + xyz position, neural shape model latent vectors), and a pressure component (None, per vertex cumulative pressure, per vertex peak pressure). All combinations of shape and pressure predictors (except None, None) were analyzed. To assess the effect of overfitting, analyses were compared using no dimensionality reduction, and principal components analysis (PCA) dimensionality reduction to 20 principal components (PCs), 50 PCs, and the rank-limited max components.

Logistic regression was fit separately for each gait in the dataset using the L-BFGS optimizer and L2-penalized logistic regression. Performance was assessed on held out data in 5-fold cross validation with 10-repeats and the mean results over repeats reported. Features were standardized; feature scaling and PCA dimensionality reduction were performed on the training folds. Cross validation was run separately for each data input (shape x pressure) and dimensionality reduction method. AUROC mean and 95% confidence interval were computed over the 10 gait trials. Dimensionality reduction changed AUROC performance for pressure metrics at most 0.03 points (full data: cumulative pressure = 0.56, peak pressure = 0.58; 50 PCs: cumulative pressure = 0.59, peak pressure = 0.60; 20 PCs: cumulative pressure = 0.58, peak pressure = 0.59). For shape latents, dimensionality reduction progressively decreased AUROC, with 20 PCs reducing it by 0.14 points, likely owing to the fact that the shape vectors are already compressed representations with a loss explicitly designed to reduce feature correlation^28^ (full data: 0.69; 50 PCs: 0.62; 20 PCs: 0.55). Results present the full raw data as inputs.

#### Cartilage Thickness Biomarker Comparisons

We extracted mean cartilage thickness in standard anatomical regions of interest as well as simulated pressure-based regions. For each knee and each gait, we identified contacting vertices (cumulative pressure > 0 Pa) on the medial femur and, separately, the medial tibia, and retained those whose cumulative pressure exceeded the 10th percentile among contacting vertices (**Extended Data Figure 5**). A vertex was assigned to that knee’s final contact region if it was retained in at least half of that knee’s gaits. The anatomically defined medial weight bearing femur region was identified using a previously developed rule-based algorithm^37^ as implemented in multiple previous investigations^28,68^(**Extended Data Figure 5**). Mean thickness was computed as the average thickness over all vertices in each defined region of interest. The outcome of interest was mean cartilage thickness change (Δ 24 month: 24-month thickness – baseline thickness, Δ 48 month: 48-month thickness – baseline thickness). Responsiveness was quantified using the standardized response mean (SRM) = mean(Δ) / SD(Δ) across knees. The 95% confidence intervals for each SRM, and for the paired difference in SRM between region definitions, were obtained by resampling knees with replacement (5,000 replicates).

**Extended Data Figure 5.**
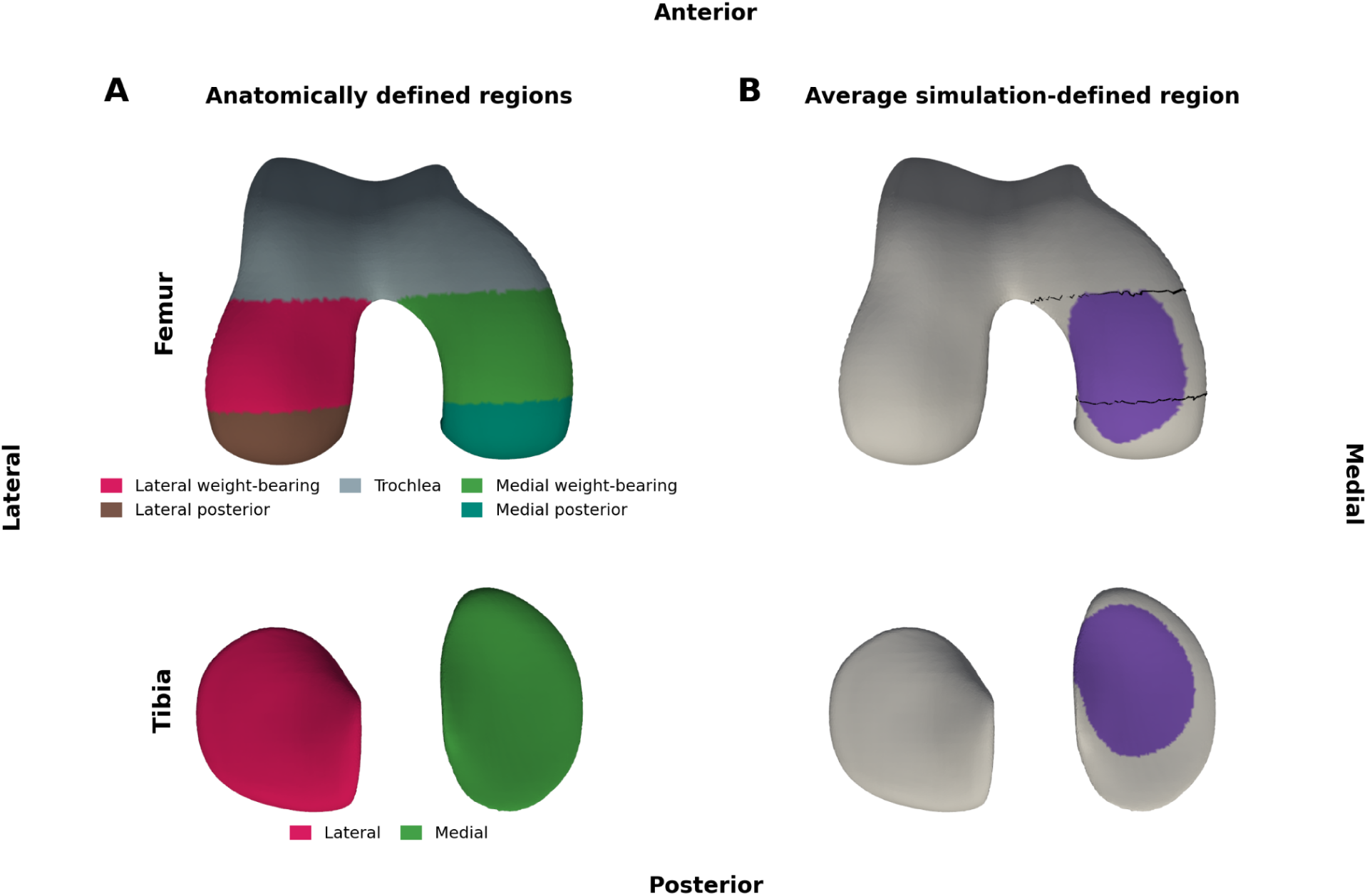
The anatomically (A) and mechanically (B) defined regions of interest used in Figure 8 and Extended Data Figure 3. **A.** Anatomical subregions on the reference cartilage surfaces. Femoral cartilage was divided anterior-to-posterior into trochlea, weight-bearing and posterior regions using the intercondylar notch (where the trochlear cartilage separates into the two condyles) as the anterior boundary and a point 60 % of the way from the notch to the most posterior femoral cartilage as the posterior boundary. Condylar cartilage was then divided medially and laterally at the notch. Tibial cartilage was divided into the medial and lateral plateaus. The standard comparators are the medial weight-bearing femur and the whole medial tibia. **B.** The average simulation-defined region. Each knee’s region comprises the medial vertices whose cumulative pressure exceeded the 10th percentile of contacting vertices in at least half of that knee’s ten gaits; the panel shows the vertices retained, on average over all knees, in at least half of the gaits (n = 750). The black outline on the femur marks the anatomical medial weight-bearing region for comparison; no outline is drawn on the tibia because its comparator is the entire medial plateau.

To determine the ability to detect between-group differences in longitudinal change, Cohen’s d was computed for incident vs. healthy and mild OA vs. healthy thinning. Ninety-five percent confidence intervals for each Cohen’s d estimate were obtained by resampling knees within each group (5,000 replicates). The difference in Cohen’s d between the simulation-defined and anatomically defined regions were computed on the same resampled knees within each replicate, yielding a paired 95% confidence interval and a two-sided bootstrap p-value.

## Data Availability

The original MRI and the healthy-subject gait data are publicly available (MRI: nda.nih.gov/oai; gait: github.com/opensim-jam-org/jam-resources). All knee MRI segmentations, 3D anatomic models and simulation results from this study will be shared in a single Hugging Face repository before peer-review publication.

https://github.com/gattia/comak_knee_oa_simulation

## Software and Data Availability

The original MRI and the healthy-subject gait data are publicly available (MRI: nda.nih.gov/oai; gait: github.com/opensim-jam-org/jam-resources). All knee MRI segmentations, 3D anatomic models and simulation results from this study are shared in a single Hugging Face repository (released on publication). The analysis code is available at github.com/gattia/comak_knee_oa_simulation, with instructions to download the data, install the libraries, re-run the simulations and statistics, and redraw every figure. The analysis code uses three custom libraries, also public and tagged paper1-v3: nsosim (github.com/gattia/nsosim), pycomak (github.com/gattia/pycomak) and the OpenSim-JAM fork of opensim-core (github.com/gattia/opensim-core). The neural shape model (github.com/gattia/nsm), mesh-processing (github.com/gattia/pymskt) and statistical-mapping (github.com/gattia/pyKneeSPM) libraries are also publicly available.

## Acknowledgements

This study would not be possible without OAI data, a public-private partnership funded by the National Institutes of Health, and private funders Merck Research Laboratories; Novartis Pharmaceuticals Corporation, GlaxoSmithKline; and Pfizer, Inc. We acknowledge research support from the Wu Tsai Human Performance Alliance, National Institutes of Health (R01AR079431, R01AR083018, R01EB002524, R01HL167974, R01HL169345, R01AR077604, P41EB027060, P50HD118632), the Department of Defense (W81XWH-18-1-0590), the Department of Veterans Affairs (I01CX002527), and ARPA-H (AY2AX000045, 1AYSAX0000024-01).

## Author Contributions

Conceptualization: A.A.G., F.K., A.S.C. Methodology and software: A.A.G., K.R.M., A.C., C.O., A.E., C.S. Formal analysis: A.A.G. Data collection: J.K., S.U. Data curation: A.A.G., M.S.W., T.T. Resources and funding acquisition: G.E.G., S.L.D., A.S.C. Original draft: A.A.G. Review & editing: all authors.

## Competing Interests

AAG is a shareholder of NeuralSeg, NodeAI, and GeminiOV. ASC is a co-founder of Cognita, holds equity interest in Cognita, Subtle Medical, LVIS Corp, and Brain Key, and provides consulting services to Patient Square Capital, Chondrometrics GmbH, and Elucid Bioimaging. The other authors declare no competing interests.

## Use of AI tools

Claude language models (Opus, Fable) were used to support software development, and to provide feedback in the writing process.

## Notes

### Clinical Trial

NCT02767570

### Author Declarations

The original MRI and the healthy-subject gait data are publicly available (MRI: nda.nih.gov/oai; gait: github.com/opensim-jam-org/jam-resources). The osteoarthritis gait data was collected as part of a clinical trial. The trial protocol was approved by the Institutional Review Board of Stanford University and the US Department of Veterans Affairs Office of Research and Development. All participants provided written informed consent. This study is registered with ClinicalTrials.gov, NCT02767570, and is closed to enrollment.

